# Cerebrospinal fluid proteomics for predictive assessment of Alzheimer’s Disease risk

**DOI:** 10.1101/2025.10.24.25337921

**Authors:** Saima Rathore, Eric B. Dammer, Anantharaman Shantaraman, Fang Wu, Duc M. Duong, Edward J. Fox, Erik C. B. Johnson, James J. Lah, Alzheimer’s Disease Neuroimaging Initiative, Nicholas T. Seyfried, Allan I. Levey

**Affiliations:** Department of Biomedical Informatics, Emory University School of Medicine, Atlanta, GA, USA; Department of Neurology, Emory University School of Medicine, Atlanta, GA, USA; Department of Biochemistry, Emory University School of Medicine, Atlanta, GA, USA; Center for Neurodegenerative Disease, Emory University School of Medicine, Atlanta, GA, USA

## Abstract

Alzheimer’s disease (AD) involves early molecular changes beyond amyloid-β (Aβ) and tau, that create heterogeneous disease biology, giving rise to variable disease initiation and highly variable longitudinal trajectories. Accurately predicting trajectories is vital for design of clinical trials and for clinical care, yet current CSF and PET biomarkers provide limited predictive capabilities despite their excellent diagnostic value. We performed CSF proteomics using tandem-mass-tag mass spectrometry in 1,104 ADNI participants with extensive longitudinal assessments. Machine learning–derived protein panels accurately predicted two classes of outcomes. First, they identified several key inflection points along the disease trajectory, including onset of 1) amyloid plaque pathology (Aβ- to Aβ+; AUC=0.88), 2) symptoms (asymptomatic to symptomatic; AUC=0.89), and 3) functional decline (MCI [due-to-AD] to AD Dementia; AUC=0.88). Second, protein panels forecast longitudinal trajectories of decline, spanning both clinical domains (cognition and function) and pathological process, including tau accumulation measured by tau-PET neocortical standardized uptake value ratio (SUVR) and neurodegeneration indexed by hippocampal volume and FDG-PET SUVR. Proteomics panels outperformed conventional CSF- and PET-based Aβ and tau markers. Importantly, these predictions were driven by novel mechanisms, spanning synaptic signaling, proteostasis, metabolic stress, vascular remodeling, and immune dysregulation, that anchor distinct inflection points and shape long-term trajectories. Together, these findings position CSF proteomics as a powerful approach for anticipating disease onset and progression, with direct implications for patient stratification and personalized intervention.

## Introduction

Alzheimer’s disease (AD), the most common age-related neurodegenerative disorder, begins with neocortical amyloid-beta (Aβ) deposition that drives tau spread and progressive neurodegeneration culminating in dementia^1–3^. These hallmark pathologies emerge years before clinical symptoms, prompting significant interest in biomarkers to identify individuals at-risk for progressive neurodegeneration^2,4,5^. This understanding led to the development of ‘A/T/N’ (Aβ/tau/neurodegeneration) biomarker framework by the National Institute on Aging and the Alzheimer’s Association^1^.

Considerable progress has been made in detecting hallmark AD pathology. Cerebrospinal fluid (CSF) Aβ42, total tau, and phosphorylated tau (pTau) have become critical tools^6^, with plasma analogs (Aβ42/Aβ40, pTau217/pTau181/pTau231) showing comparable accuracy^7,8^. The microtubule-binding region has also emerged as an excellent marker for tau tangles, correlating strongly with tau-PET^9^. Despite these advances, current biomarkers capture only part of AD’s complex biology and offer limited prognostic value in forecasting key disease-related events and long-term trajectories. Even tau, though modestly predictive of progression in several studies^10–12^, including our own^11,13^, provides an incomplete view of disease dynamics. This gap constrains progress in two key ways. First, it limits our ability to monitor longitudinal changes and evaluate therapeutic effects. Second, it impacts clinical trials, fundamentally targeting distinct inflection points: primary prevention enrolls Aβ-negative at-risk individuals (e.g., Dominantly Inherited Alzheimer Disease; DIAN-TU^14^) to delay conversion to Aβ-positivity; secondary prevention enrolls asymptomatic Aβ-positive individuals (e.g., Anti-Amyloid Treatment in Asymptomatic AD; A4^15^, AHEAD 3-45^16^) to delay symptom onset; and symptomatic trials aim to slow progression from mild cognitive impairment (MCI) to AD Dementia. All trial types face challenges with cohort enrichment, as many participants remain stable within study windows, reducing statistical power to detect treatment effects. Identifying individuals most likely to convert or decline rapidly would enable more efficient trial design, smaller sample sizes, and greater sensitivity to therapeutic benefit.

Proteomic research complements genomic and transcriptomic data, exemplified by Accelerating Medicine Partnership for AD (AMP-AD) efforts to identify biomarkers and therapeutic targets. In prior work, we applied mass spectrometry (MS)-based proteomics to ∼2,000 AMP-AD brain and CSF samples^17–23^, developed a targeted selection reaction monitoring assay to quantify 48 AD-linked proteins in CSF^24^, and used data from ADNI and Emory Healthy Brain Study to build machine learning (ML) models that refined AD staging, predicted disease trajectories, and identified Aβ-independent proteins linked to cerebral amyloid angiopathy^25–27^. Collectively, these studies showed that proteomic markers are clinically translatable and capture mechanisms beyond Aβ and tau.

Building on these findings, we generated a deep CSF proteome (2,492 proteins) using high-resolution tandem-mass-tag (TMT)-MS to capture diverse pathophysiological processes in 1,104 ADNI participants (**Fig. S1-S2; Table. S1-S2**). The multi-modal ADNI dataset allowed integrated predictive modeling using machine learning across proteomic, imaging, and clinical domains. Leveraging this framework, we identified proteomic panels that predicted (i) key inflection points, including pathologic conversion (Aβ- to Aβ+), symptomatic onset (asymptomatic to symptomatic MCI [due-to-AD] or AD Dementia), and clinical progression (MCI [due-to-AD] to AD Dementia), and (ii) surpassed conventional biomarkers in predicting longitudinal decline in cognition, function, and neuroimaging measures. Pathway analyses supported a staged model of AD pathophysiology, in which distinct yet overlapping biological mechanisms govern disease trajectories. Longitudinal event modeling further mapped the sequence of biological changes preceding symptom onset. Together, these findings define molecular signatures of onset and progression, offering a framework for precision trial enrollment and improved monitoring of AD trajectories.

## Results

### Proteomic biomarkers accurately predict onset of AD pathology

Aβ deposition begins silently one to two decades before symptom onset, marking the earliest detectable phase of AD biology. Primary prevention of AD therefore aims to intervene before this pathological conversion, making its accurate prediction essential for effective strategies. To this end, we performed high-resolution TMT-MS based quantitative proteomics on CSF samples from 1,104 ADNI participants (**Fig. S1-S2; Table. S1-S2**), enabling deep proteome coverage with high precision^29,30^. In total, 102,544 peptides were identified, mapping to 3,907 protein groups, of which 2,492 proteins were quantified in at-least 50% of samples and included in downstream analyses. Participants were first classified by AT status using the CSF Aβ42/pTau181 ratio (cutoff=39.20 ^28^) into AT- (n=552; controls and biomarker-confirmed non-AD) and AT+ (n=552; biomarker- confirmed AD) groups. Among AT- participants, clinically diagnosed subgroups included healthy controls (n=277), non-AD MCI (n=259), and non-AD Dementia (n=16); among AT+ participants, subgroups included asymptomatic AD (n=100), MCI [due-to-AD] (n=304), and AD Dementia (n=148).

To predict onset of AD pathology, we analyzed 309 participants who were Aβ-negative by both the Aβ42/pTau181 ratio^28^ and [^18^F] AV45 PET, and who had longitudinal PET scans available to determine whether they subsequently converted to Aβ-positivity. Among these individuals, 271 remained Aβ-negative (Aβ-) over an average follow-up of 4.85 years, while 48 converted to Aβ- positivity (cAβ+) within an average of 4.34 years.

Differential abundance analysis revealed 40 proteins increased and 33 decreased (P<0.05) in converters, marking early molecular changes associated with the onset of Aβ pathology **(Fig. 2a; Table. S3; Fig. S3)**. Notably, BMP7, known to protect against Aβ-induced neurotoxicity^31^, may be upregulated as an early neuroprotective response to rising Aβ levels. Similarly, ANGPT2, which promotes Aβ-induced angiogenesis and blood-brain barrier (BBB) disruption^32^, likely reflects early vascular stress in response to emerging pathology. In converters, protective proteins such as RAB7A, which facilitates Aβ clearance through autophagy/lysosomal pathways^33^, and LRIG1, involved in synaptic maintenance, were reduced, potentially contributing to disease onset^34^. AChE, the enzyme that hydrolyzes acetylcholine at synaptic clefts^35^, thereby terminating cholinergic transmission, was elevated in Aβ- individuals, suggesting a protective role against conversion to Aβ+. Importantly, these participants were not receiving AChE inhibitor therapy, indicating that the elevation was endogenous rather than treatment induced.

**Fig. 1.**
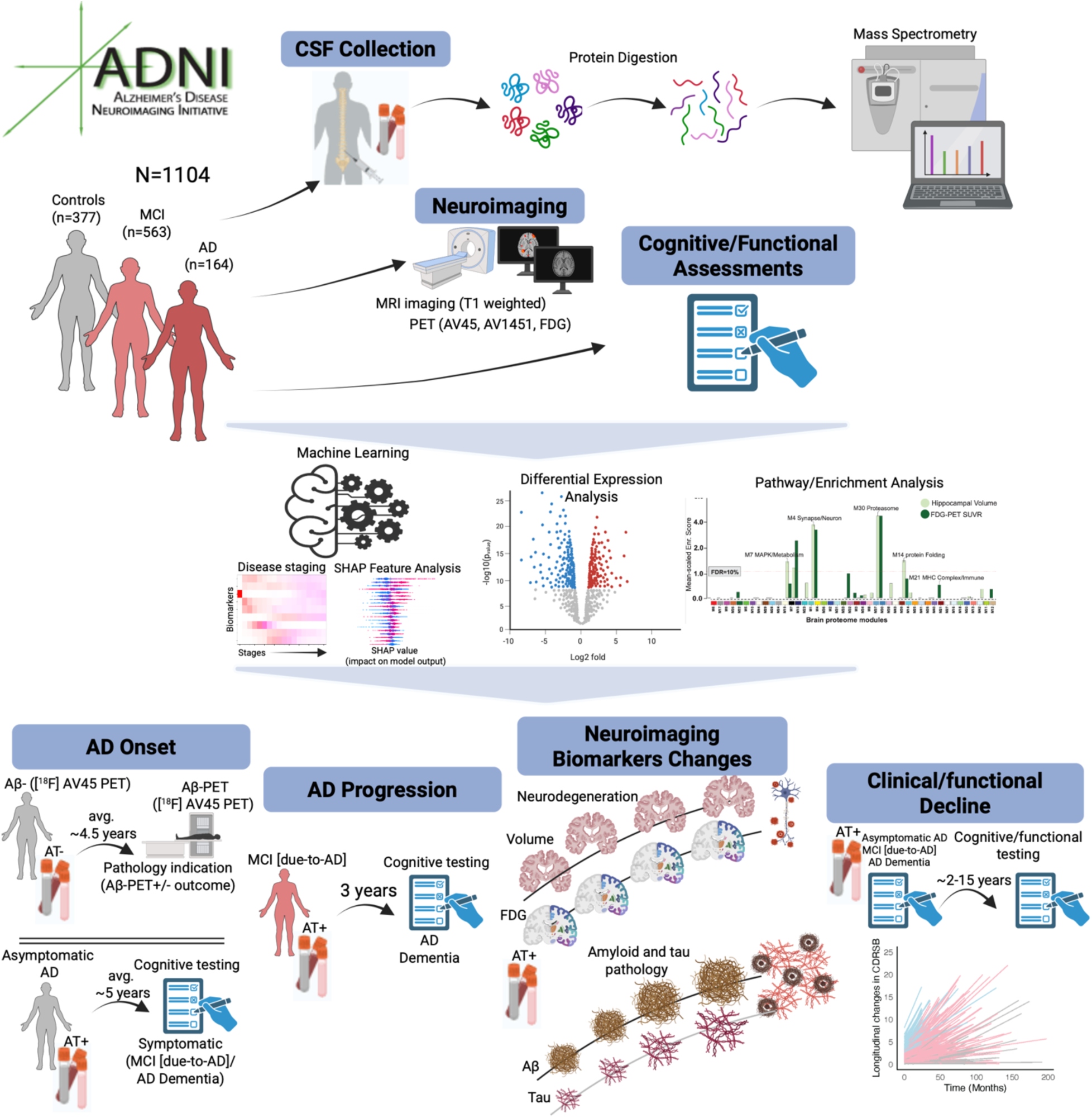
Multimodal proteomic, imaging, and clinical data integration identifies proteins associated with onset and progression in AD. Schematic overview of the study design. CSF proteomic profiles from 377 controls [277 AT-, 100 AT+], 563 MCI [259 AT-, 304 AT+] and 164 Dementia [16 AT-, 148 AT+] participants as defined by their clinical diagnosis and CSF Aβ42/pTau181 ratio (39.20 cutoff^28^) were analyzed. AT-participants, including controls, MCI and Dementia, are referred as controls, non-AD MCI, and non-AD Dementia, and AT+ participants (i.e., biomarker-confirmed AD), including controls, MCI and Dementia, are referred as asymptomatic AD, MCI [due-to-AD], and AD Dementia. Differentially abundant proteins across these outcomes were identified and mapped to pathways using Gene Ontology analysis. These proteins were subsequently mapped to brain-derived networks and assessed for enrichment within network modules from prior work^17^. Machine learning models trained on these proteins identified highly predictive CSF protein panels for pathological and symptomatic onset in AD, as well as for progression from MCI [due-to-AD] to AD Dementia. Using longitudinal clinical outcomes, predictive protein panels were derived for clinical progression in biomarker-confirmed AD participants. Finally, proteo-radiomic analysis of biomarker-confirmed AD participants with both baseline CSF and longitudinal neuroimaging data revealed highly predictive protein panels of changes in tau accumulation, neurodegeneration, and metabolic dysfunction.

**Fig. 2:**
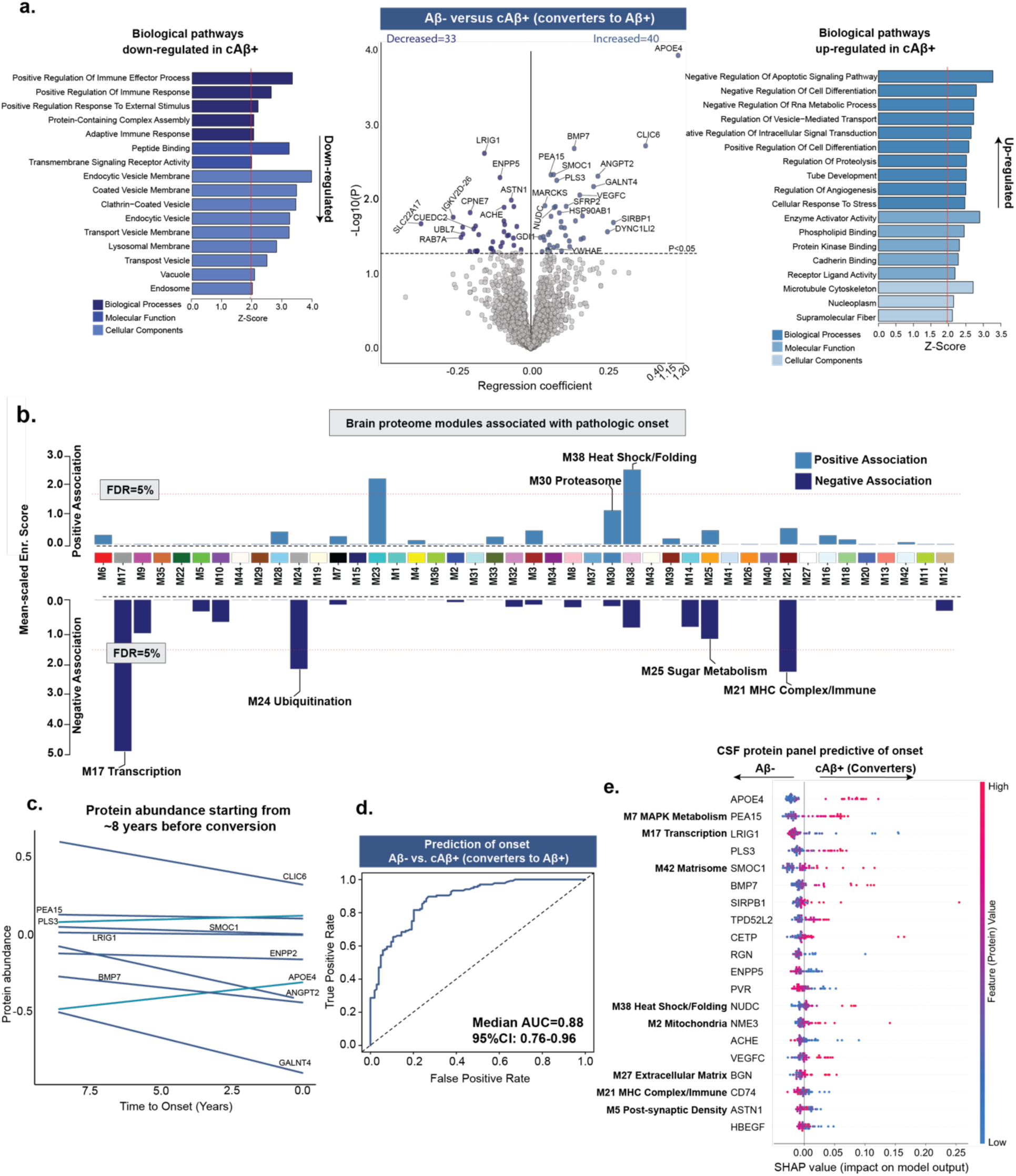
CSF proteomes predict onset of AD pathology. Participants with baseline CSF measures and longitudinal Aβ-PET ([^18^F] AV45) scans were leveraged to predict the onset of Aβ pathology. ***a,*** Volcano plot shows differentially abundant proteins when comparing Aβ- participants who remained Aβ- [Aβ-, n=271] versus those who converted to Aβ+ status [cAβ+, n=48], as determined on follow-up [^18^F] AV45 PET scan. The black line represents the threshold of P<0.05, above which proteins were considered significant. For clarity, top-ranking proteins are labeled. Gene ontology analysis was performed to identify biological processes associated with the conversion from Aβ- to Aβ+ using the proteins significantly associated with conversion, as shown in the volcano. Upregulated and downregulated pathways are separately shown to highlight the directionality of the associations. ***b,*** CSF proteins associated with pathologic onset were mapped to previously defined brain proteomic network modules^17^. The horizontal red dotted line indicates the threshold for permutation test false discovery rate (FDRs of 5%) above which the enrichment of proteins correlated with pathologic onset was considered significant. Modules M17, M24, M38, and M21, showing significant enrichment, and additional relevant modules are labeled. Unlabeled but significant modules lacked defined annotations in the original network study. ***c,*** Population-level protein abundance in cAβ+ participants plotted relative to time-to-conversion, defined as the interval between CSF collection and first Aβ-PET positivity. The top ten proteins (ranked by P value) are shown. ***d,*** Proteomics data were used to train classification model to distinguish Aβ-from cAβ+. LASSO based feature selection along with random forest classification-based model were trained. ROC curves from 100 permuted runs illustrate the median AUC based on an 80-20% train-test split for Aβ-from cAβ+ (AUC=0.88 [0.76-0.96]). Higher AUC values indicate better classification performance, with values closer to 1.0 reflecting greater sensitivity and specificity. ***e,*** SHapley Additive exPlanations (SHAP) based feature analysis is conducted on the model’s outcome to determine contributions of different proteins in predictive modelling. Top SHAP feature contributions corresponding to the median AUC in panel ***(d)*** are shown. Each row represents a protein, and each dot reflects an individual’s data. The x-axis shows the SHAP value, indicating how strongly each protein influenced the model’s prediction. Dot color reflects protein abundance (red = high, blue = low). Proteins are ranked by overall impact, with APOE4, PEA15, LRIG1, PLS3, and SMOC1 among the most influential. Brain module names are shown for CSF proteins present in the brain proteome and assigned to network modules.

Gene Ontology (GO) biological-process enrichment indicated mechanisms associated with the onset of Aβ pathology, including increased activity of processes mediating suppression of apoptotic signaling and cell differentiation, repression of RNA metabolic activity, and tighter regulation of vesicle-mediated trafficking and intracellular signal transduction (**Fig. 2a**). Notably, converters also showed broad downregulation of both innate and adaptive immune programs, including reduced expression of microglial/vascular (CLIC1, PECAM1, FCGBP) and antigen-presentation (HLA-B, HLA-DRA, HLA-DRB1, CD74) pathways, in addition to marked downregulation of endo-lysosomal traffic pathways. To contextualize CSF signatures of converters in terms of brain pathology, we mapped these proteins onto brain co-expression modules (M1-M44) from prefrontal cortex proteomic networks in our prior study^17^. Proteins showing increased abundance in converters were enriched in proteases (M30) and heat-shock (M38) modules, whereas proteins showing decreased abundance clustered in modules related to metabolism (M25), transcription (M17), ubiquitination (M24), heat-shock (M38), and MHC complex/immune (M21) (**Fig. 2b; Table. S4**). This suggests an early immune-quiescence state, where proteostatic defenses (cell-intrinsic stress defenses) are upregulated but coordinated immune surveillance is muted, potentially allowing Aβ pathology to accumulate silently. Overall, these processes suggest that the brain is already attempting to adapt to emerging pathology, mobilizing stress and defense responses even before overt amyloid deposition.

Differential abundance analysis identified proteins that differed significantly between converters and non-converters, but their temporal dynamics during Aβ conversion remain unclear. To investigate this, protein abundance levels from 36 converters were plotted against time-to-conversion (**Fig. 2c**), defined as the time-interval between CSF sample collection and the first Aβ- PET positive scan. Because each participant contributed only a single proteomic measurement, these curves represent population-level trends rather than within-person trajectories. Within this framework, APOE4 levels were higher at earlier time relative to conversion, suggesting that increases occur years before detectable Aβ accumulation, whereas ANGPT2, BMP7, CLIC6 (endosomal-vesicular/receptor trafficking), and GALNT4 (O-glycosylation of secretory/receptor proteins), though elevated in converters, showed declining abundance approaching conversion, consistent with an early compensatory response that diminishes as pathology consolidates.

To identify a protein panel predictive of Aβ pathology onset, we trained random forest machine learning model^36^ along with least absolute shrinkage and selection operator (LASSO)^37^, resulting in a median AUC of 0.88 (95% CI:0.76-0.96) in distinguishing cAβ+ and sAβ- across 100 permuted runs while splitting the data in an 80-20 train/test configuration (**Fig. 2d; Table. S5**). To identify proteins driving model predictions, we applied SHapley Additive exPlanations (SHAP) analysis^38^, which revealed that the protein panel most predictive of Aβ onset (**Fig. 2e)** included APOE4 and other proteins involved in metabolism (PEA15), transcription (LRIG1), matrisome (SMOC1), heat shock/folding (NUDC), mitochondria (NME3), extracellular matrix remodeling (BGN), MHC complex/immune function (CD74), post-synaptic biology (ASTN1), and many more. These proteins individually were not always the most differentially expressed, yet collectively these proteins as a panel are an excellent predictor of the future onset of AD pathology and the biological processes that are intimately involved in the initiation of disease.

### Proteomic biomarkers predict transition from preclinical to symptomatic AD

Beyond Aβ conversion, symptom onset marks a second key inflection point with considerable individual variability. Identifying those who will develop symptoms is critical for secondary prevention and for advancing therapies. Large prevention initiatives such as A4^15^ and A3-45^16^ in Aβ+ cognitively unimpaired adults illustrate these efforts and highlight the challenges of targeting individuals before clinical onset. To test whether proteomic signatures could capture the earliest shifts toward clinical disease, we analyzed CSF proteome from 100 asymptomatic AD participants followed longitudinally. Of these, 64 remained stable (sAsym; mean follow-up=5.28-years) and 36 converted to MCI [due-to-AD] or AD Dementia (cAsym; mean conversion=4.59-years).

Differential abundance analysis identified 151 proteins increased and 135 decreased in converters (P<0.05; **Fig. 3a; Table. S6**). Most of the proteins with decreased abundance (TYRO3, CACNA2D3, THY1, EPHA5, NELL2, NPY, PCSK1, FAM174A) were linked to neuronal and axonal development, synaptic structure, and ephrin signaling (**Fig. 3b**), with enrichment in pre- and post- synaptic compartments, indicating broad synaptic disruption. In contrast, upregulated processes involved immune and vascular pathways, including adaptive and innate immune responses, complement activation, fibrin clot formation, and lipid transport/metabolism, exemplified by SERPINA1, ABI3BP, F10, LAMA3, AZGP1, and IGHV6-1. Interestingly, both death-receptor signaling, marking activation of pro-apoptotic^39^ and inflammatory cascades, and negative regulation of endothelial apoptosis, suggesting activation of vascular survival mechanisms preserving BBB integrity^40^, were upregulated, revealing a dynamic tension between cell-death and pro-survival programs at symptom onset.

**Fig. 3:**
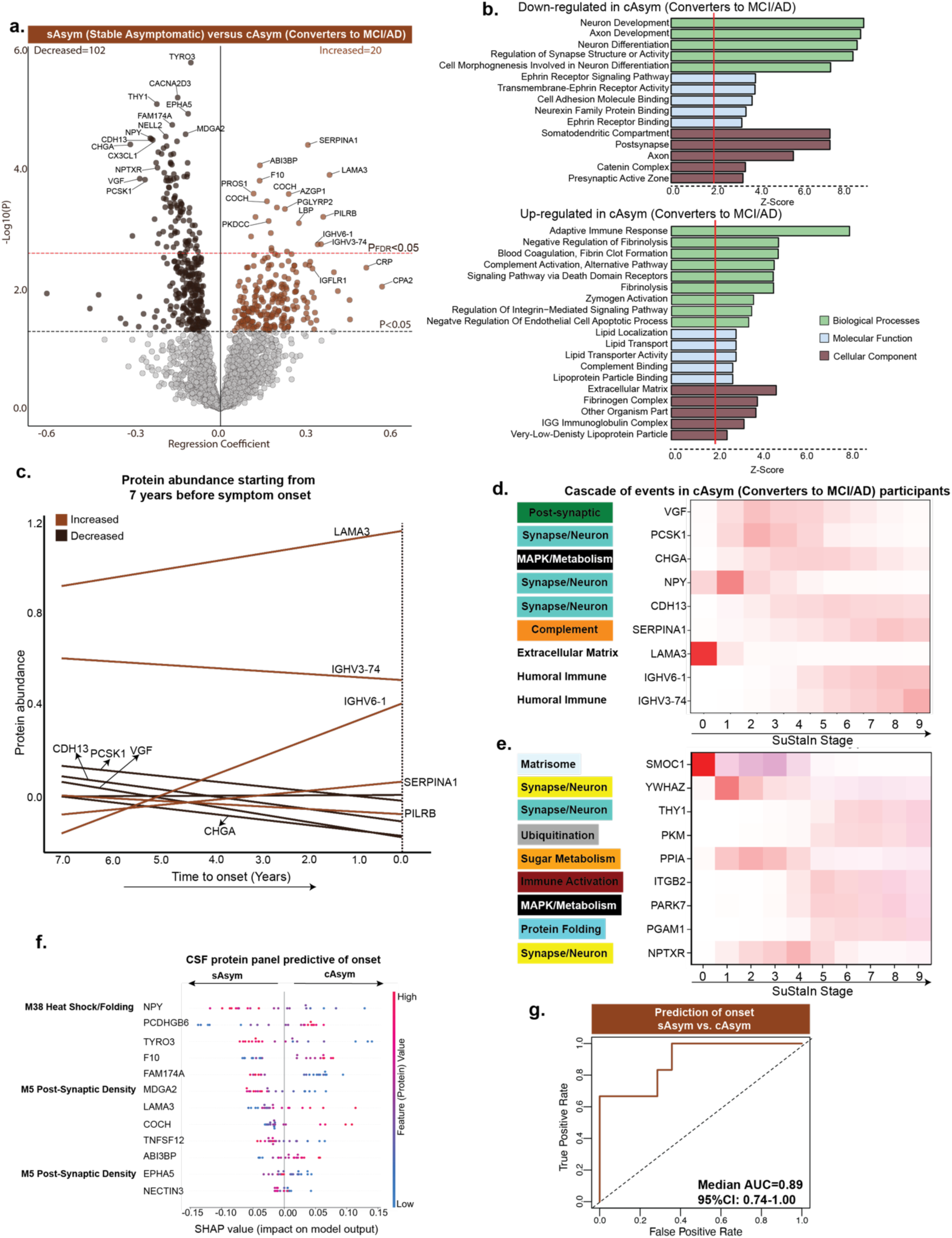
CSF proteomes predict onset of clinical symptoms. ***a,*** Volcano plot showing differentially abundant proteins between asymptomatic AD participants who remained stable (sAsym, n=64) and those who progressed to MCI [due-to-AD] or AD dementia (cAsym, n=36). Significant proteins (P<0.05; increased, n=151; decreased, n=135) are highlighted, with top proteins labeled for legibility. Black line indicates the significance threshold of P<0.05, and the red line marks the FDR-corrected threshold of 0.05**. *b,*** Functional enrichment analysis of the associations in **(a)** using GO databases, with representative summary terms is shown. ***c,*** Protein abundance in 36 cAsym participants plotted relative to time to symptom onset, measured from the date of CSF collection to first diagnosis of MCI [due-to-AD] or AD Dementia. Top-most 10 proteins are selected based on p-values. ***d,*** Event-based modeling with SuStaIn clustering using the top proteins identified in **(a)** in 36 cAsym participants, illustrating the sequence of proteomic alterations during progression from asymptomatic to symptomatic stages. ***e,*** Event-based modeling of 36 cAsym participants using the proteins identified in the DIAN-TU cohort^42^ to be altered in autosomal dominant AD before onset. ***f,*** Proteomics data were used to train a classification model to distinguish cAsym from sAsym. LASSO-based feature selection along with a random forest classification-based model was trained. Top SHAP feature contributions to the model outcome are shown for cAsym versus sAsym. Each row represents a protein, and each dot reflects an individual’s data. The x-axis shows the SHAP value, indicating how strongly each protein influenced the model’s prediction. Dot color reflects protein abundance (red = high, blue = low). Proteins are ranked by overall impact, with NPY, PCDHGB6, and TYRO3 among the most influential. Brain module names are shown for CSF proteins present in the brain proteome and assigned to network modules. ***g,*** Median AUC (AUC = 0.89 [0.74-1.00]) is shown when the model was tested across 100 permuted runs based on an 80-20% train-test split for asymptomatic converters versus non-converters. Higher AUC values indicate better classification performance, with values closer to 1.0 reflecting greater sensitivity and specificity.

To extend the findings from differential abundance analysis, we mapped the significant CSF proteins onto our previously defined brain proteomic network^17^ (**Fig. S4; Table. S7**) to find shared molecular modules. We identified four brain modules enriched for CSF proteins conferring a negative association with symptom onset: M5 (post-synaptic), M7 (MAPK/metabolism- intracellular lipid use/synthesis), M15 (synaptic vesicles) and M1 (synapse/neuron); and one brain module, M26 (complement/acute phase), conferring a positive association with symptom onset, reinforcing the pathways in **Fig. 3b**. Together, these changes highlight a shift from synaptic and neuronal integrity toward extracellular matrix remodeling (ECM), inflammation, immune activation, vascular dysregulation, and metabolic stress during clinical conversion.

To further resolve proteins associated with symptom onset (**Fig. 3a**), we aligned abundance levels of proteins with time-to-conversion (**Fig. 3c**) and fitted regression lines across 36 converters. This analysis revealed early increases in LAMA3, IGHV6-1, and SERPINA1, contrasted by declines in CDH13, PCSK1, VGF, and CHGA, with both patterns detectable up to 7 years before conversion, the maximum conversion window captured in our data (**Fig. 3c**). These opposing patterns highlight coordinated shifts across various pathways preceding symptom onset. To define temporal ordering of pathways being altered, we applied Subtype and Stage Inference (SuStaIn) clustering^41^, which uncovered a cascade initiating with ECM remodeling (LAMA3), progressing through synaptic and neuropeptide dysfunction (VGF, PCSK1, CHGA), advancing to innate and adaptive immune activation (SERPINA1, LBP, IGHV6-1, IGHV3-74), and culminating in synaptic collapse (GABBR2) (**Fig. 3d**). Notably, a parallel trajectory we reported in the DIAN-TU study using targeted MS^42^ recapitulated this sequence, beginning with matrix disruption (SMOC1), followed by synaptic vulnerability (YWHAZ, NPTXR), then metabolic stress (PPIA, PARK7, PGAM1, PKM), transitioning to immune activation (ITGB2), and ultimately converging on synaptic deterioration (THY1) (**Fig. 3e**).

Building on these pathway-level alterations, we next examined whether proteomic shifts, modeled across all quantified proteins rather than only those reaching statistical significance, could be leveraged for prediction of progression. When distinguishing sAsym from cAsym using LASSO-based protein selection and random forest based classification as described earlier, we identified a predictive 12-protein panel (median AUC=0.89, 95% CI:0.74-1.00 across 100 runs; **Fig. 3f-g; Table. S8**) spanning axon guidance and synaptic organization (EPHA5, TYRO3, NPY, PCDHGB6), neuronal differentiation (THY1, MDGA2, NECTIN3), and vascular-immune processes including coagulation and ECM remodeling (F10, LAMA3, COCH, TNFSF12, ABI3BP), capturing the coordinated loss of synaptic integrity alongside activation of inflammatory and vascular cascades during clinical conversion.

### Proteomic signatures forecast progression to AD Dementia in MCI [due-to-AD] participants

Having established that different proteomic profiles predict disease onset, we next tested whether proteomics could also forecast progression from MCI [due-to-AD] to AD Dementia. Although most individuals eventually convert, their trajectories vary widely, with some individuals remaining stable for years, whereas others converting to Dementia rapidly^43^. Using longitudinal cognitive data from participants diagnosed with MCI [due-to-AD] at baseline CSF collection and a prespecified 3-year conversion window, we derived an analytic cohort of 186 individuals, comprising stable participants (sMCI; n=61), defined as persistent MCI beyond 3-years, and converters (cMCI; n=125), who progressed to AD Dementia within 3-years.

Relative to sMCI, converters showed increases in 111 and decreases in 223 proteins (P<0.05; **Fig. 4a; Table. S9**). These alterations reflected system under stress, with heightened activity in glycolysis, coagulation, fatty-acid metabolism, wound healing, and TOR signaling (**Fig. 4b**), reflected in the positively associated proteins (YWHAZ, YWHAE, UCHL1, ENO1, PEA15, PDXP, ACHE, FABP3, PPP3CA). Conversely, pathways essential for neuronal development and synaptic organization, including axonogenesis, axon guidance, and postsynaptic density, showed marked downregulation (**Fig. 4b**), represented by negatively-associated proteins (PTPRN2, PNOC, EPHA5, VGF, CDH8, BDNF, NPTXR, NPTX2, SCG2, PCDH, CX3CL1), indicating that as disease progresses to AD Dementia, biological resources increasingly divert toward compensatory survival mechanisms, while synaptic and neuronal processes progressively weaken. When mapped onto brain proteome modules, these proteins showed broad enrichment across synaptic and postsynaptic, heat-shock, protein folding, proteasome, and metabolic pathways, reflecting widespread dysregulation and stress responses underlying neurodegeneration, consistent with advanced disease transition to AD Dementia (**Fig. S5; Table. S10**).

**Fig. 4:**
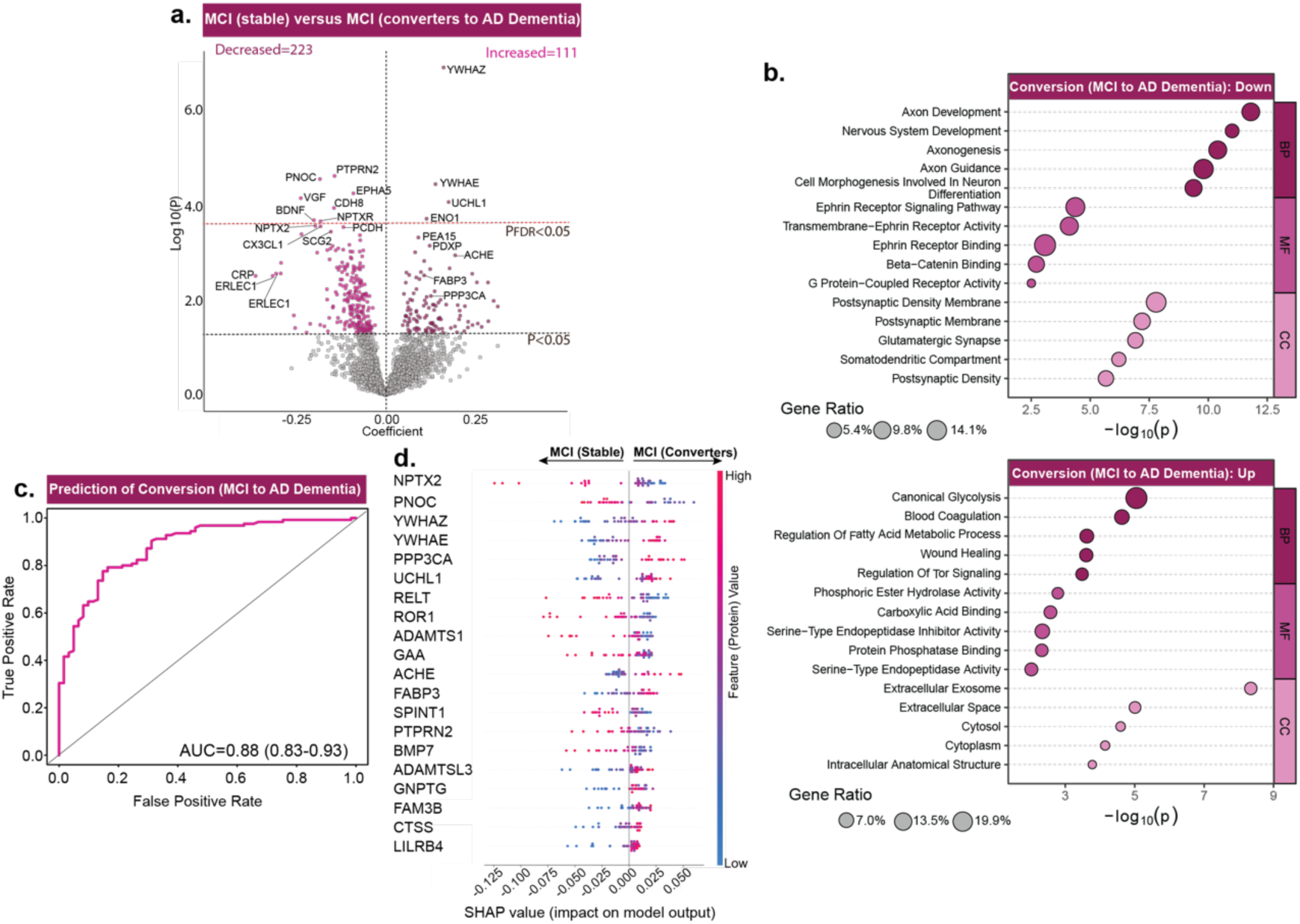
CSF proteomes predict disease progression from MCI [due-to-AD] to AD Dementia. ***a,*** Volcano plot shows differentially abundant proteins when comparing MCI [due-to-AD] participants who maintained diagnosis over 3 years [sMCI, n=61] versus those who converted to AD Dementia [cMCI, n=125]. Black line indicates the significance threshold of P<0.05, and the red line marks the FDR-corrected threshold of 0.05. Only the top proteins are labeled for legibility on the volcano plots. ***b,*** Summary terms from functional enrichment analyses using GO databases from the associations shown in (**a)**. ***c,*** Proteomics data were used to train classification model to distinguish cMCI from sMCI. LASSO-based feature selection along with random forest-based classification model were trained. ROC curves from 100 permuted runs illustrate the median Area Under the Curve (AUC) based on an 80-20% train-test split for converters from non-converters (AUC = 0.88, 95% CI: 0.83-0.93). Higher AUC values indicate better classification performance, with values closer to 1.0 reflecting greater sensitivity and specificity. ***d,*** SHAP based feature analysis is conducted on the model’s outcome to determine contributions of different proteins in predictive modelling. Top SHAP feature contributions corresponding to the median AUC model in panel **(c)** are shown for cMCI versus sMCI. Each row represents a protein, and each dot reflects an individual’s data. The x-axis shows the SHAP value, indicating how strongly each protein influenced the model’s prediction. Dot color reflects protein abundance (red = high, blue = low). Proteins are ranked by overall impact, with NPTX2, PNOC, and YWHAZ among the most influential.

Building on these pathway-level alterations, we next asked whether the proteomic shifts could enable predictive modeling of progression to AD Dementia. A random forest-based machine learning model along with LASSO-based protein selection, as explained earlier, achieved robust discrimination between cMCI and sMCI (AUC=0.88, 95% CI:0.83-0.93; **Fig. 4c**; **Table. S11**). As expected, the most predictive proteins spanned a wide swath of AD biology, capturing synaptic (NPTX2, PNOC, YWHAZ, YWHAE, PPP3CA), presynaptic (UCHL1), ECM (ADAMTS1, ADAMTSL3), neuronal-signaling (BMP7), neuroinflammatory (RELT, CTSS, LILRB4), and metabolic (FABP3, GAA, SPINT1, GNPTG, FAM3B) processes, highlighting that progression is foreshadowed by coordinated disruption across multiple molecular systems (**Fig. 4d**).

### Proteomic signatures capture biological processes that forecast rate of progressive cognitive decline and dementia severity

The biological cascade underlying AD emerges well before clinical onset and advances through asymptomatic, MCI, and Dementia stages along heterogeneous trajectories, with some individuals experiencing slow progression, whereas others declining rapidly^44,45^. This variability often dilutes treatment effects in clinical trials, rendering them statistically non-significant even when subsets of participants may benefit. Identifying the biological processes that predispose individuals to faster deterioration is essential for trial enrichment^46^ and personalized interventions. To address this, we turned to finer-grained longitudinal measures of cognition (ADAS-Cog11) and dementia severity (CDR-SB) collected over extended follow-up (**Fig. 5a**), asking whether there are biological processes that distinguish the pace of decline.

**Fig. 5.**
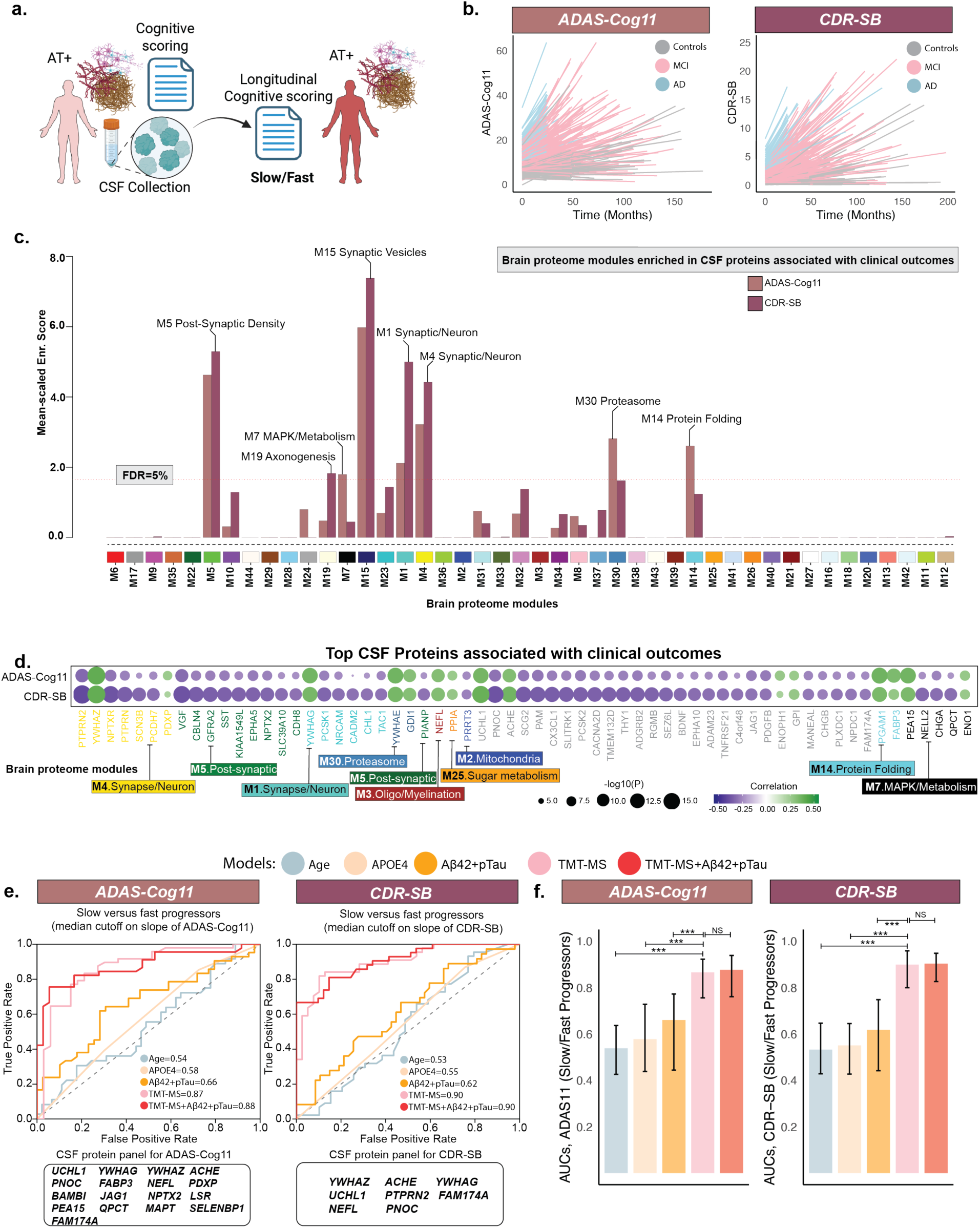
CSF proteomics predicts longitudinal trajectories of clinical and dementia severity measures beyond conventional biomarkers. ***a**,* Schematic of the overall design, where participants with longitudinal clinical outcome are selected for analysis. ***b**,* Longitudinal trajectories of participants in terms of cognitive (ADAS-Cog11) and dementia severity (CDR-SB) measures calculated using the linear mixed-effects models, which estimate both population-level (fixed) effects and subject-specific (random) slopes. Participants having at-least 3 timepoints and data available for at least 24 months were used to calculate longitudinal trajectories. ***c**,* Proteins associated with progression mapped to brain protein network modules^17^. The horizontal red dotted line indicates the threshold for permutation test false discovery rate (FDRs of 5%) above which the enrichment of proteins correlated with progression was considered significant. Labeled modules showed significant enrichment of proteins linked to progression. ***d**,* A heatmap of Pearson correlations is shown between protein abundance and rate of change in clinical outcomes. The proteins are labeled as their respective gene symbols, and the strength and direction of correlation is shown by the blue to green color scale. The top 30 proteins with the strongest correlations were selected for each clinical outcome, and their union is shown in the heatmap. Colors are assigned to individual CSF protein names based on their assignment in the brain-derived modules^17^, which are shown for proteins present in the brain proteome and assigned to network modules. ***e**,* Participants were divided into fast-versus stable/slow progressors based on median cutoff slope of each clinical outcome. ROC curves show the classification accuracy in predicting slow- and fast-progressors using the following predictors: 1) Age alone, 2) APOE with χ4 allele frequency modelled as 0,1, and 2, 3) CSF Aβ42 and pTau181 as two features (“Aβ42+pTau”), 4) the proteomics alone (“TMT-MS”), and 5) the proteomics and CSF Aβ42 and pTau181 (“TMT-MS+ Aβ42+pTau”). Random forest-based classification models were trained separately for each predictor type using an 80-20% train-test split across 100 permuted runs. LASSO-based feature selection was applied only in model no. 4, and the same set of selected proteins was combined with Aβ42 and pTau181 in model no. 5. ***f**,* Bar plots show the distribution of AUCs across the 100 permuted runs of each classification model. Permutations were kept the same across all the models to perform a fair performance comparison. Sample sizes were: ADAS-Cog11 (N = 406), and CDR-SB (N = 414). (*p<0.05, **p<0.01, ***p<0.001). NS (non-significant).

Linear mixed-effects (lmer) models were employed to estimate rates-of-change in ADAS- Cog11 and CDR-SB, including participants with longitudinal data spanning ≥2-years and at least ≥3 timepoints (**Fig. 5b**). Consistent with established synaptic underpinnings of cognitive decline^47^, brain-module overlap showed that modules centered on synaptic vesicle cycling, neuronal communication, and postsynaptic density were associated with the rate-of-change in both the measures (**Fig. 5c; Table. S13**). Beyond the synaptic modules, association strengths diverged: stress-response and proteostasis modules, including MAPK/metabolism (M7), proteasome (M30), and protein folding (M14), were stronger for ADAS-Cog11, consistent with its sensitivity to early cognitive decline, whereas modules reflecting structural disconnection and circuit-level remodeling, including axonogenesis (M19), were stronger for CDR-SB, aligning with its emphasis on later functional impairment (**Fig. 5c**). This asymmetry reflects both biological staging and instrument sensitivity, highlighting complementary but non-mutually exclusive processes captured by each measure.

To further dissect these drivers, a proteome-wide association identified proteins linked with clinical decline (ADAS-Cog11: Increased=49, Decreased=170; CDR-SB: Increased=104, Decreased=261; PFDR<0.01; **Fig. S6; Table. S12**). A heatmap of the top-30 proteins illustrate how individual signals (proteins) cluster within broader biological pathways (**Fig. 5d**). Proteins such as YWHAZ, YWHAG, NPTXR, NPTX2, VGF, SST, and PCSK1 were consistently associated with progression across both the measures, underscoring their central role in mediating clinical decline. Other significant proteins localized to proteasome (GDI1, YWHAE), protein-folding (PGAM1, FABP3), metabolism (QPCT, CHGA, NELL2), and axonogenesis. Together, these findings suggest that ADAS-Cog11 and CDR-SB capture complementary dimensions of AD biology, with differential sensitivity to staging, yet converging on shared molecular drivers of decline (**Fig. S10**).

To identify protein panels predictive of progression, individuals were classified as slow or fast progressors by median cutoff, and machine learning models were trained. Highly predictive proteomic panels were identified, distinguishing progressor groups with high accuracy (median AUC: ADAS-Cog11=0.87, CDR-SB=0.90; **Fig.5e-f; Table. S14-S15**). For ADAS-Cog11, predictive proteomic panel spanned synaptic and axonal biology, neuronal signaling, and metabolic/stress-response pathways (**Fig. 5e**; UCHL1, YWHAZ, ACHE, YWHAG, PDXP, FABP3, NEFL, NPTX2, LSR, QPCT, PEA15, SELENBP1, MAPT, FAM174A, PNOC), while an overlapping subset (YWHAZ, YWHAG, UCHL1, FAM174A, NEFL, PNOC), with the addition of ACHE and PTPRN2, best predicted CDR-SB progression **(Fig. 5e)**. By contrast, canonical predictors (Age, CSF based Aβ42 and pTau181, and APOE4), long recognized as limited in forecasting clinical decline, showed only modest performance (AUCs∼0.53-0.66; **Fig. 5e-f**) and adding them to proteomics offered little gain (median AUC: ADAS-Cog11=0.88; CDR-SB=0.90), suggesting that proteomics capture variance beyond conventional biomarkers. Importantly, proteomic models also outperformed tau-PET-based predictions, currently considered gold-standard^12,48^, in a subset of participants with both datatypes (proteomics and tau-PET) available (**Fig. S7**). Together, these findings across two clinical endpoints show that conventional biomarkers reflect susceptibility, whereas proteomic signatures more directly capture the molecular biology of decline, potentially involving networks that drive tau spread, synaptic loss, and metabolic stress, positioning proteomics as a powerful tool for risk stratification and trial enrichment in AD.

### Longitudinal neurodegenerative biomarker changes are modeled more accurately with proteomic profiling than with conventional measures

Having established associations with clinical decline, we next turned to neurodegenerative imaging markers, which provide sensitive readouts of brain structure and metabolism. Among the most widely used measures, FDG-PET reflects cerebral glucose metabolism and structural MRI- derived hippocampal volume indexes atrophy, together representing the ‘N’ component of A/T/N framework^1^. Although these markers provide readout of progression^49^, they do not directly uncover the molecular mechanisms that drive them. To bridge this gap, we examined associations between proteomic profiles and FDG-PET standardized uptake value ratio (SUVR) and MRI- derived hippocampal volume.

Using repeated measures of hippocampal volume (normalized by intracranial volume) and FDG-PET SUVR [Average of angular, temporal, and posterior cingulate regions], we applied linear mixed-effects models to estimate subject-specific rates of neurodegenerative change (**Fig.6b**). Processes associated with neurodegeneration (**Fig. 6d**) overlapped with synaptic and metabolic modules linked to clinical decline (**Fig. 5c-d**) but also extended to sugar metabolism (PPIA), MAPK/metabolism (PEA15, ENO1), proteasome (GDI1, YWHAE), M1-cell interaction (PGAM2), protein-folding (FABP3, PGAM1), oligodendrocyte/myelination (NEFL), mitochondrial function (CDH6), and synaptic processes (YWHAZ, YWHAG, YWHAB) (**Fig. 6d,f; Fig. S8; Table. S16-S17**). As metabolic changes often precede atrophy, FDG-PET provided a sensitive window into early dysfunction, uniquely implicating pathways related to synaptic vesicle cycling and neurotransmission (SV2B, PODXL, BDNF), mitochondrial dynamics (FIS1, RBP3), lysosomal/glycoprotein catabolism (MAN2B1, INAFM2), immune-metabolic signaling (FAM19A5, BTN2A1, LY6E), and neurotrophic-inflammatory balance (FST, BDNF), capturing early metabolic and synaptic vulnerability preceding structural degeneration.

**Fig. 6.**
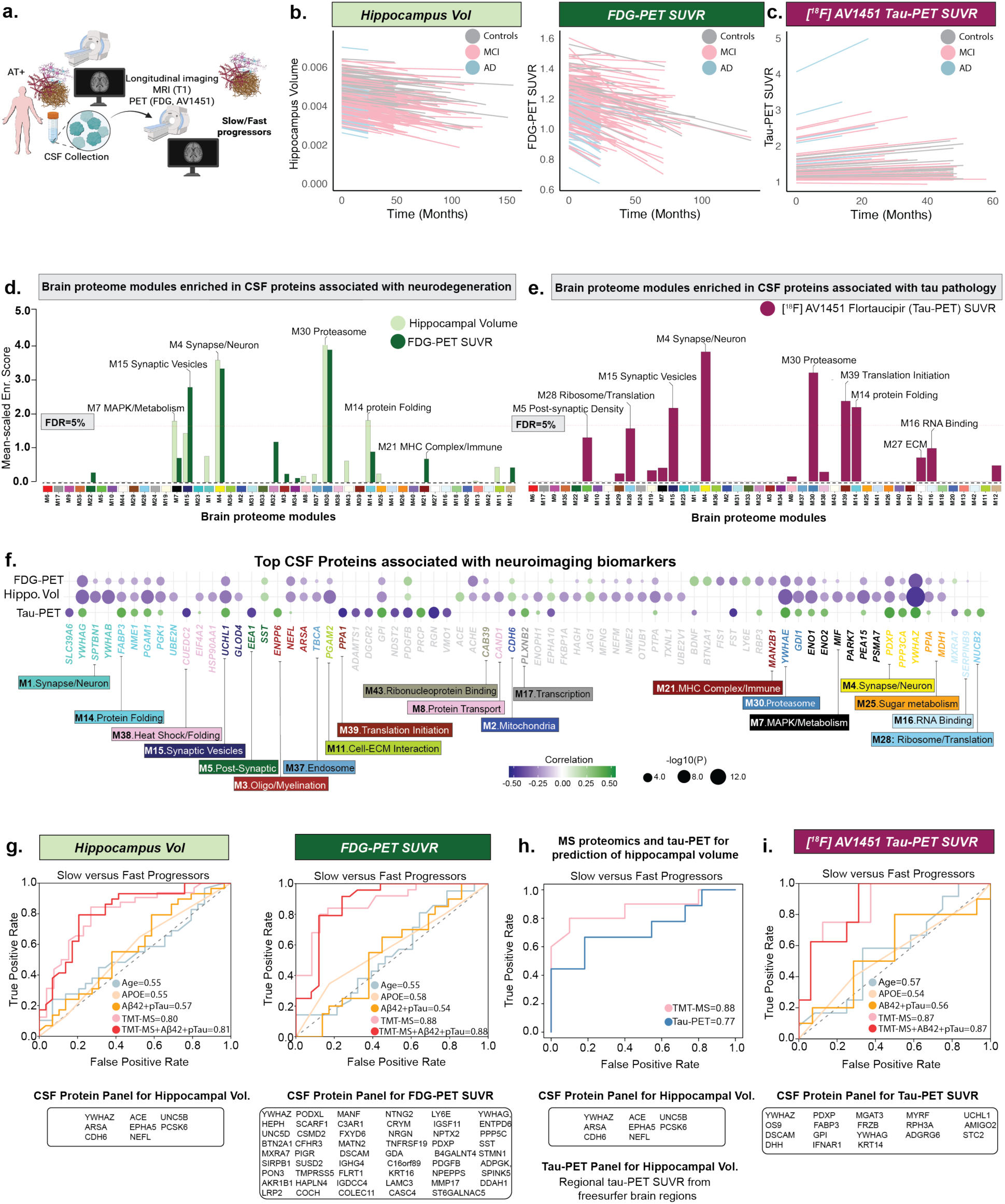
CSF proteomics predicts longitudinal changes in neurodegeneration and pathology accumulation. ***a-c,*** Longitudinal trajectories of participants (i.e., rate of change) in terms of neurodegeneration (hippocampal volume, and FDG-PET SUVR; average of angular, temporal, and posterior cingulate regions) and pathology accumulation (neocortical tau-PET SUVR; [^18^F] AV1451) measures calculated using the linear mixed-effects models. Participants having at-least 2 timepoints and data available for at least 12 months were used to calculate longitudinal trajectories, except for hippocampal volume where sufficient data was available to restrict the analysis to 3 timepoints and for at least 24 months. ***d-e,*** Proteins associated with rate of change were mapped to brain protein modules^17^. The horizontal red dotted line indicates the threshold for permutation test false discovery rate (FDR of 5%) above which the enrichment of proteins correlated with rate of change was considered significant. Modules labeled include both statistically significant and biologically relevant non-significant modules, highlighting pathways implicated in progression. ***f,*** A heatmap of Pearson correlations is shown between protein abundance and rate of change in neuroimaging biomarkers. The proteins are labeled as their respective gene symbols, and the strength and direction of correlation is shown by the blue to green color scale. The top 30 proteins with the strongest correlations were selected from each category, and their union is displayed in the heatmaps. Colors are assigned to individual CSF protein names based on their assignment in the brain-derived modules^17^, which are shown for proteins present in the brain proteome and assigned to network modules. ***g,*** Participants were stratified into fast and slow progressors based on the median slope of each neurodegeneration biomarker. ROC curves show the classification performance for predicting progressor groups using five predictor sets: 1) Age alone, 2) APOE4 genotype with χ4 allele dosage coded as 0, 1, and 2, 3) CSF Aβ42 and pTau181 as two features (“Aβ42+pTau”), 4) proteomics alone (“TMT-MS”), and 5) combined proteomics and CSF Aβ42 and pTau181 (“TMT-MS+ Aβ42+pTau”). Random forest classifiers were trained separately for each predictor type using an 80-20% train-test split across 100 permuted runs. LASSO-based feature selection was applied only to model 4, and the same set of selected proteins was combined with Aβ42 and pTau181 in model 5. Identical permutation splits were maintained across all models for each outcome to ensure fair performance comparison. ***h,*** Mean tau-PET SUVR values from FreeSurfer-defined neocortical regions were used, in addition to proteomics, to predict rate of change in hippocampal volume in participants with both proteomics and tau-PET data available. LASSO-based feature selection and model training were performed separately for the proteomic and tau-PET predictors. ***i,*** Following the same framework as in panel **g,** five predictor models were trained to predict the rate of change in neocortical tau-PET SUVR. Sample sizes were: hippocampal volume (N = 290), FDG-PET SUVR (N = 241), neocortical tau-PET SUVR (N = 83). NS (non-significant).

Conventional biomarkers provide limited insight into neurodegeneration rates and weakly predict clinical trajectories; accordingly, we evaluated whether proteomic profiles could stratify progression more effectively. Participants were classified as slow and fast progressors by median slope, and machine learning models identified protein panels with predictive value beyond Aβ and tau. The proteomic panels achieved strong discrimination (hippocampal volume: AUC=0.80; FDG-PET SUVR: AUC=0.88; **Fig. 6g; Tables. S18-19**), outperforming conventional biomarkers with minimal gain from adding Aβ42 and pTau181. Importantly, proteomic biomarkers also outperformed tau-PET–based measures in predicting hippocampal atrophy (**Fig. 6h**). Together, these findings suggest that proteomic profiles capture variance beyond conventional CSF and imaging biomarkers, consistent with the view that neurodegeneration reflects the interplay of multiple molecular networks rather than single-pathway readouts.

### Proteomic data reveal pathway-level perturbations that improve forecasting of tau accumulation beyond established biomarkers

Neuroimaging and pathological studies demonstrate that cortical tau colocalizes with cortical atrophy and predicts cognitive decline^50,51^, with emerging evidence also indicating that tau deposition within specific brain regions drives domain-specific cognitive impairments^13,52^. These observations have established tau as a central therapeutic target for disease-modifying interventions^53,54^ and key biomarker in interventional trials^55–57^. Although tau progression has been modelled using neuroimaging and clinical data^10,11^, yet the biological pathways that drive these changes remain elusive. To this end, we evaluated proteomic profiles as predictors of neocortical tau-PET SUVR trajectories, thereby probing the biological processes underlying tau pathology. Leveraging longitudinal [¹⁸F] AV1451 PET data, we estimated subject-specific rates of neocortical SUVR change (**Fig. 6c**).

Enrichment analysis revealed that many tau-associated proteins clustered within synaptic and post-synaptic modules (**Fig. 6e**), overlapping with those linked to clinical progression (**Fig. 5c-d**) and neurodegeneration (**Fig. 6d**), indicating shared molecular architecture across outcomes. Among modules related to protein homeostasis, the core proteostasis modules M14 (protein-folding and metabolism) and M30 (proteasome) were broadly altered across all stages of the disease, underscoring their central role in AD pathophysiology (**Fig. 2-4**). Beyond these canonical proteostasis processes, tau-associated proteins further enriched RNA-binding (M16), translation-initiation (M39), and ribosomal/translation (M28) modules, suggesting that tau pathology extends proteostatic stress to RNA metabolism and protein-synthesis machinery, compounding impairments in folding and clearance pathways (**Fig. 6e**) in parallel with clinical and neuroimaging decline.

To further dissect these proteomic signatures, we performed differential abundance analysis that identified 138 proteins positively and 151 negatively associated with tau accumulation (P<0.05; **Fig. S9; Table. S20**). A heatmap of the top-ranked proteins illustrates their organization into broader biological pathways (**Fig.6f**). Regucalcin (RGN), a Ca²⁺-homeostasis regulator that supports PMCA/SERCA function and dampens Ca²⁺-dependent kinases, phosphatases, and calpains^58^, showed one of the strongest associations, implicating calcium-handling stress in tau pathology. Proteostasis-related hits included PPIA/Cyclophilin A (peptidyl-prolyl isomerase/chaperone)^59^, MXRA7 (matrix-remodeling glycoprotein), SERPINB9 (serine-protease inhibitor with cytoprotective/immune functions)^60^, and NUCB2 (Golgi/secreted Ca²⁺-binding protein)^61^. Additional proteins linked to ECM remodeling (ADAMTS1), metabolism (PRCP, PPA1), heparan-sulfate modification (NDST2), proteasomal degradation (CUEDC2), myelin/choline metabolism (ENPP6), vascular adhesion (DGCR2), and oxidative stress responses (VMO1) (**Fig. 6f; Table. S20-S21**). Collectively, these data and GO analysis (**Fig. S9**) position tau-PET progression at a proteostasis-synaptic-endolysosomal nexus, with dysregulated calcium handling, translational control, and protein-quality control emerging as convergent drivers of tau-related neurodegeneration.

We next asked whether proteomic profiles could anticipate which participants would accumulate tau faster. Using the median tau-PET neocortical SUVR slope as a cutoff, proteomic data discriminated fast from slow progressors with high accuracy (median AUC=0.87 across 100 permuted runs; **Fig.6i; Table. S22**), outperforming conventional biomarkers. These findings indicate that proteomic signatures capture the systems-level consequences of tau pathology, providing a more integrative framework for predicting disease trajectories.

## Discussion

Our study establishes an unbiased CSF proteomic framework that captures molecular signatures underlying onset and progression in AD. Integration with brain proteome^17^ refined the molecular architecture, revealing coordinated shifts across distinct biological pathways. Further, incorporation of ADNI CSF profiles with machine learning yielded predictive and prognostic proteomic panels, validated through rigorous independent permutation testing reinforcing the generalizability and translational potential of these panels. Together, these findings show that onset and progression in AD are governed by distinct yet interacting mechanisms, underscoring the need to disentangle these processes for targeted intervention.

To disentangle these intertwined mechanisms, we examined the molecular and clinical inflection points that punctuate disease evolution, revealing how AD pathology emerges and progresses across stages. Protein-based integrated framework identified individuals at risk before pathological conversion, redefining AD risk assessment. By identifying Aβ- individuals predisposed to amyloid accumulation or cognitively normal individuals approaching conversion, these models could offer a minimally invasive, scalable first-line screen. Clinically, they could guide confirmatory testing, support monitoring, and enable timely intervention. In primary prevention, early identification of biologically at-risk individuals creates opportunities to delay or even prevent onset through lifestyle or pharmacologic strategies^62^. In secondary prevention, prospective enrichment of participants most likely to convert within a trial window increases power and reduces costs, accelerating evaluation of disease-modifying therapies^63,64^. With further validation, protein-based prediction could anchor precision prevention across both preclinical and symptomatic stages, pinpointing molecular inflection points where disease trajectories diverge and therapeutic leverage is greatest^58^.

The biological pathways governing transitions across inflection points reveal a staged architecture of AD pathophysiology. The earliest phase, marked by Aβ onset, reflects a shift from a regenerative, synaptic plasticity state toward stress, repair, and maladaptive degeneration. Metabolic strain, vascular and inflammatory repair responses, and chronic stress emerge while neuronal growth and connectivity decline, echoing longitudinal and single-cell observations^65,66^. Proteins upregulated at this stage reinforce these processes: BMP (compensatory response to Aβ toxicity)^67^, CLIC6 (endosomal-lysosomal/synaptic vulnerability)^68^, ANGPT2 and VEGFC (angiogenesis, vascular remodeling, BBB disruption), and APOE4 and SMOC1 (neurovascular dysfunction and ECM remodeling). As symptoms arise, compensatory synaptic and neurodevelopmental mechanisms collapse, and destructive immune-vascular cascades take over, exemplified by TYRO3, NPTXR, VGF, SERPINA1, ABI3BP, and LAMA3. Neurons lose resilience, while adaptive immunity, coagulation, complement activation, and apoptosis reshape the tissue environment, marking the critical switch from silent pathology to cognitive impairment. Finally, in progression to AD Dementia, synaptic and axonal programs are further suppressed, while glycolysis, coagulation, fatty-acid metabolism, and TOR signaling are upregulated, suggesting compensatory metabolic and vascular remodeling in the context of advancing neuronal decline. Together, this sequence delineates how early immune quiescence permits pathology onset, how symptom emergence is driven by synaptic collapse with immune-vascular activation, and how conversion to Dementia reflects the convergence of neuronal degeneration with maladaptive metabolic compensation.

Another important finding of the study is elevated acetylcholinesterase (AChE) levels in Aβ- individuals who remained pathology-free (**Fig. 2a**), which suggests that preserved cholinergic tone may confer resilience in preclinical AD stages. This aligns with evidence linking anticholinergic drug use to greater Aβ burden, worse cognition, and increased dementia-risk^69,70^. Together with variable cognitive outcomes in Aβ-targeting trials^57,71^, these findings imply that baseline cholinergic tone may influence therapeutic response and that early cholinergic dysfunction may require combined Aβ-neurotransmitter interventions. AChE expression was also significantly higher in clinically diagnosed AD/MCI participants receiving cholinesterase inhibitors compared with untreated individuals (P=1.16×10^-9^; P<0.0001 after adjusting for diagnosis). These distinct patterns suggest dual origins of AChE elevation, pharmacologic upregulation in symptomatic stages versus endogenous preservation of cholinergic tone in resilient, Aβ- individuals.

Our framework goes beyond predicting disease onset to delineating longitudinal trajectories that capture the pace and heterogeneity of progression in AD. While prior proteomics studies have linked disease phenotypes to proteins^72–74^, they rarely predict individualized trajectories, limiting precision medicine applications. Here, we not only identified proteomic panels predictive of longitudinal trajectories but also demonstrated the mechanics underlying variable trajectories, thereby offering a dynamic view of progression at the individual-level. These proteomic panels provided strong prognostic value, surpassing A/T/N biomarkers and even tau-PET based measures (**Fig. 5-6**; **Fig. S7**), currently considered the most accurate predictors of longitudinal AD trajectories^11,48^. This suggests that CSF protein signatures capture broader and potentially earlier disease processes than regional tau accumulation. In interventional trials, prospective enrichment of participants most likely to decline rapidly will increase statistical power and reduce costs, accelerating the evaluation of disease-modifying therapies^63,64^.

The biological underpinnings of clinical decline converged on pathways reflecting metabolism and energy regulation, protection against cellular stress, preservation of synaptic and structural integrity required for neuronal signaling, and proteasomal clearance of damaged proteins (**Fig. 5**). A key finding was the strong, positive association between PEA15 and clinical outcomes. PEA15 is implicated in the clearance of Aβ plaques by astrocytes, glial cells that play a role in brain health, and their ability to clear Aβ is important in preventing the buildup of plaques^75^. Research also suggests that PEA15 could be a potential biomarker for AD, especially in the preclinical stages^76^. Notably, many of these same pathways were also implicated in neurodegenerative markers, especially hippocampal volume, indicating a strong overlap between the molecular drivers of cognition and those of structural decline. FDG-PET was enriched for early metabolic, mitochondrial, and synaptic pathways, reflecting their sensitivity to functional vulnerability before overt pathology. By contrast, tau-PET mapped onto later programs: translation initiation, driving protein synthesis; transcriptional regulation, reshaping neuronal and immune gene networks; protein folding and heat-shock responses, protecting proteostasis but when impaired fueling toxic aggregation; and ECM remodeling, maintaining structural support but when dysregulated disrupting synaptic and vascular integrity. RGN, a calcium-homeostasis regulator, emerged as one of the strongest predictors of longitudinal tau accumulation, implicating early failure of neuronal calcium buffering in the cascade toward tau hyperphosphorylation and aggregation. This finding aligns with experimental evidence that regucalcin protects against Aβ toxicity and that calcium dysregulation accelerates tau pathology, positioning RGN as a novel link between disrupted calcium handling and tau progression^77,78^. The developed tau panel can sensitively detect later pathology as a cost-effective PET alternative. Together, they support trial stratification, optimize anti-Aβ therapy outcomes given the established relationship between anti- Aβ treatment efficacy and tau burden^57,71^, and serve as pharmacodynamic markers of target engagement and efficacy.

In summary, these findings define how AD pathology can be anticipated and tracked across distinct stages of disease evolution through proteomic profiling. Pathological onset marks the earliest shift, when compensatory defenses fail and amyloid accumulates silently. Symptom onset reflects collapse of synaptic and neurodevelopmental programs alongside immune-vascular activation and cognitive decline. Third, prediction for conversion to AD Dementia reflects the progression from early symptomatic impairment to dementia, characterized by consolidation of neuronal degeneration and accompanying metabolic and vascular remodeling. Finally, distinguishing slow from fast progressors through proteomic signatures enables patient stratification after onset, when multitarget therapies may slow decline. Together, these proteome-derived stages highlight four actionable windows for intervention spanning preclinical to symptomatic AD.

Our study has several limitations. First, we did not test whether CSF-derived signatures can be recapitulated in plasma, which would enable minimally invasive screening. Second, we included all available participants for predicting onset without restricting the observation period. Finally, our analyses relied exclusively on the TMT-MS platform, leaving open the possibility of platform-specific bias. In future work, we will extend these analyses to NuLISA-based assays in plasma samples from a larger ADNI cohort. This expanded effort will enable orthogonal validation across platforms and matrices, directly test the translatability of CSF-derived signatures to blood, and address the current limitations related to cohort size and measurement bias. Together, these future directions aim to strengthen and generalize our current findings, which already demonstrate that CSF-based proteomics captures broader AD-related mechanisms and improves prediction of disease onset and progression beyond conventional biomarkers.

## Materials and methods

### ADNI patient cohort characteristics

Data used in the preparation of this article were obtained from the Alzheimer’s Disease Neuroimaging Initiative (ADNI) database (adni.loni.usc.edu). The ADNI was launched in 2003 as a public-private partnership, led by Principal Investigator Michael W. Weiner, MD. The primary goal of ADNI has been to test whether serial magnetic resonance imaging (MRI), positron emission tomography (PET), other biological markers, and clinical and neuropsychological assessment can be combined to measure the progression of mild cognitive impairment (MCI) and early Alzheimer’s disease (AD).

The ADNI study was approved by the Institutional Review Boards at each participating ADNI site (see full list here: http://adni.loni.usc.edu). All procedures were performed in accordance with relevant guidelines and regulations, and informed consent was obtained from all subjects prior to enrollment. The current study was approved by the ADNI Data and Publications Committee (ADNI DPC). Data used in the present study were downloaded on Oct 20th, 2024, and inclusion is thus restricted to subjects whose data was uploaded to the ADNI database prior to this date.

Participant recruitment for ADNI is approved by the Institutional Review Board of each participating site. All ADNI participants undergo standardized diagnostic assessment that renders a clinical diagnosis of either control, MCI, or AD using standard research criteria. Control participants had no subjective memory complaints, tested normally on Logical Memory II of Weschler Memory Scale, had an MMSE between 24-30, and a CDR of 0 with memory box score of 0. A subject is diagnosed as MCI if the study participant (i) reports concern due to impaired memory function; (ii) obtains a Mini Mental State Examination (MMSE) score between 24 and 30; (iii) a Clinical Dementia Rating Scale (CDR) score of 0.5; (iv) a score lower than expected (adjusted for years of education) on the Wechsler Memory Scale Logical Memory II (WMS-II); and (v) reports preserved function of daily living. AD participants also exhibited subjective memory concerns but also met NINCDS/ARDA criteria for probable AD. AD participants also showed abnormal memory function on Logical Memory II subscale from the Weschler Memory Scale, an MMSE of 20-26, and CDR of 0.5 or 0.1.

This study was designed to identify whether CSF proteomics can facilitate diagnosis, predict onset of the disease, and estimate longitudinal disease trajectory. To test the diagnostic and prognostic utility of CSF proteomics and compare that to existing AD biomarkers, CSF collected at baseline visits were assayed from 1,104 participants recruited from ADNI. Samples were randomized and blinded for mass-spectrometry (MS) proteomics analyses. Inclusion criteria for the current study were enrollment in ADNI1/GO or ADNI2/3, an available baseline CSF sample, and longitudinal cognitive outcome, clinical records, and imaging measurements.

### CSF markers of Aβ42 and pTau181

CSF Aβ42 and pTau181 were measured using the Elecsys immunoassays (Roche Diagnostics). A pre-established cutoff of 39.20 on the CSF Aβ42/pTau181 ratio was used to define AD biomarker positivity^28^. CSF was chosen over PET to create the AT categories to maximize the sample size, because a large percentage of participants did not undergo Aβ-PET or tau-PET in the ADNI cohorts. Participants below the cutoff were considered AT- (n=552; negative ratio reflecting participants without a neurodegenerative disease diagnosis) and the ones above the threshold were considered as AT+ (n=552; positive ratio, reflecting participants with AD pathology). Controls, MCI and Dementia in AT-group are referred as Controls, non-AD MCI, and non-AD Dementia, and the corresponding participants in AT+ group are referred as asymptomatic AD, MCI (due-to-AD), and AD Dementia.

### CSF samples preparation and mass spectrometry

#### Isobaric Tandem Mass Tag (TMT) peptide labeling

Each sample was re-suspended in 100 mM TEAB buffer (50 μL). The TMT labeling reagents (5mg; TMTpro^TM^ 16-plex Label Reagent, Lot: # YA357799; 134C and 135N lot# YB370079, ThermoFisher Scientific) were equilibrated to room temperature, and anhydrous ACN (200 μL) was added to each reagent channel. Each channel was gently vortexed for 5 min, and then 10 μL from each TMT channel was transferred to the peptide solutions and allowed to incubate for 1 h at room temperature. The reaction was quenched with 5% (vol/vol) hydroxylamine (5 μl) (Pierce). All channels were then combined and dried by SpeedVac (LabConco). The combined sample was then resuspended in 500 uL of 0.1% TFA and then diluted 1:1 with 4% H3PO4 and desalted with a 30 mg MCX column (Waters). Samples were loaded onto the MCX column and then washed with 100 mM ammonium formate in 2% formic acid. This is followed by a methanol wash and finally the samples were eluted with 300uL of 5% ammonium hydroxide in methanol. The eluates were then dried to completeness using a SpeedVac (LabConco).

### High-pH off-line fractionation

Dried samples were re-suspended in high pH loading buffer (0.07% vol/vol NH4OH, 0.045% vol/vol FA, 2% vol/vol ACN) and loaded onto a Water’s BEH 1.7 um 2.1mm by 150mm. A Thermo Vanquish was used to carry out the fractionation. Solvent A consisted of 0.0175% (vol/vol) NH4OH, 0.01125% (vol/vol) FA, and 2% (vol/vol) ACN; solvent B consisted of 0.0175% (vol/vol) NH4OH, 0.01125% (vol/vol) FA, and 90% (vol/vol) ACN. The sample elution was performed over a 25 min gradient with a flow rate of 0.6 mL/min. A total of 192 individual equal volume fractions were collected across the gradient and subsequently pooled by concatenation into 96 fractions and dried to completeness using a SpeedVac.

### Liquid chromatography tandem mass spectrometry

All fractions were resuspended in an equal volume of loading buffer (0.1% FA, 0.03% TFA, 1% ACN) and analyzed by liquid chromatography coupled to tandem mass spectrometry. Peptide eluents were separated on Water’s CSH column (1.7um resin 150um by 15 cm) by a Vanquish Neo (ThermoFisher Scientific). Buffer A was water with 0.1% (vol/vol) formic acid, and buffer B was 99.9% (vol/vol) acetonitrile in water with 0.1% (vol/vol) formic acid. The gradient was from 3% to 35% solvent B over 17 mins followed by column wash and equilibration for a total of 23 mins. Peptides were monitored on a Orbitrap Astral spectrometer (ThermoFisher Scientific) fitted with a high-field asymmetric waveform ion mobility spectrometry (FAIMS Pro) ion mobility source (ThermoFisher Scientific). Two compensation voltages (CV) of -45 and -60 were chosen for the FAIMS. Each cycle consisted of one full scan acquisition (MS1) with an m/z range of 400-1500 at 120,000 resolution and standard settings and as many tandem (MS/MS) scans in 1.5 seconds. The Astral higher energy collision-induced dissociation (HCD) tandem scans were collected at 35% collision energy with an isolation of 0.5 m/z, an AGC setting of 100%, and a maximum injection time of 20ms. Dynamic exclusion was set to exclude previously sequenced peaks for 30 seconds within a 5-ppm tolerance window.

### Database search

6958 raw files (across 65 TMT-18 plexes; deposited at www.synapse.org with the SynID: syn59804727) were searched using FragPipe (version 22.0), as described above. The FragPipe pipeline relies on MSFragger (version 4.0)^79,80^ Percolator^81^ for peptide identification, MSBooster^82^ and Percolator^81^ for false discovery rate (FDR) filtering and downstream processing. The search was performed with a database of canonical Human proteins downloaded from Uniprot (20,402; accessed 02/11/2019), as well as sequences for specific peptides for APOE4 and 2 alleles^83^ alongside ABETA40 and 42 peptides as described^83^ (total of 20,405 sequences). The workflow used in FragPipe followed default TMT-16 plex parameters, used for both TMT-16 and TMT-18 experimental design. Briefly, precursor mass tolerance was -20 to 20 ppm, fragment mass tolerance of 20 ppm, mass calibration and parameter optimization were selected, and isotope error was set to -1/0/1/2/3. Enzyme specificity was set to semi-tryptic and up to two missing trypsin cleavages were allowed. Peptide length was allowed to range from 7 to 42 and peptide mass from either 200 to 5,000 Da. Variable modifications that were allowed in our search included: oxidation on methionine, N-terminal acetylation on protein, N-terminal acetylation on peptide along with off-target TMT tag modification on Serine, Threonine and Histidine, with a maximum of 3 variable modifications per peptide. Peptide Spectral Matches were validated using Percolator^81^. The FDR threshold was set to 1% and protein and peptide abundances were quantified using Philosopher for downstream analysis. Before performing any abundance analysis, the data from all batches were merged and protein levels were first scaled by dividing each protein intensity by intensity sum of all proteins in each sample followed by multiplying by the maximum protein intensity sum across all samples. Instances where the intensity was ‘0’ were treated as ‘missing values’. The searched data matrix has been deposited with ADNI Biofluid Biomarker Core.

### Tunable approach for median polish of ratio (TAMPOR)

We used TAMPOR (Tunable Approach for Median Polish of Ratio) to remove technical batch variance in the proteomic data, as previously described^84^. TAMPOR removes inter-batch variance while preserving variance caused by biological changes in the protein abundance values, normalizing to the median of selected samples.

### Regression of unwanted covariates

The protein abundance matrices are then subjected to non-parametric bootstrap regression by subtracting the trait of interest (batch of the sample) multiplied by the median estimated coefficient of fitting for each protein in the log2(abundance) matrix.

### Neuroimage data processing

#### Structural MRI processing

Each subject underwent a standardized 3T MR imaging protocol. Imaging sequences included T2* gradient recalled-echo, T1-weighted 3D MPRAGE, and FLAIR sequences (http://adni.loni.usc.edu/methods/documents/mri-protocols/). Before any subject was scanned using this protocol, an ADNI phantom was used to assess linear and nonlinear spatial distortion, signal-to-noise ratio, and image contrast, which were reviewed by a single quality-control center to ensure harmonization among the sites.

### [^18^F] Florbetapir AV45 and [^18^F] Flortaucipir AV1451 image processing

This study includes Aβ ([^18^F] Florbetapir AV45) PET, tau ([^18^F] Flortaucipir AV1451) PET, and structural MRI (T1-weighted) scans obtained through the ADNI database^85,86^. Preprocessed images from ADNI, with realigned frames, head position corrected through linear transformation, standardized voxel size, and smoothed to a uniform resolution of 6mm, were used in this study. Aβ-PET scans were analyzed in each participant’s native space, using their structural MRIs acquired closest to the Aβ-PET scan. The structural MRIs were segmented into cortical regions of interest and reference regions for each subject using FreeSurfer. The Aβ-PET data were then realigned, and the mean of all frames was used to co-register the Aβ-PET data with the corresponding structural MRI.

For each subject and time point, cortical standardized uptake value ratio (SUVR) images were generated by dividing voxel-wise Aβ-PET uptake by the average uptake from the whole cerebellum reference region. Aβ-PET SUVR values were quantified within each FreeSurfer Desikan-Killiany region (left, right, and volume-weighted bilateral)^87^. For tau-PET images, a similar procedure was followed. The tau-PET images were realigned, and the mean of all frames was used to co-register the tau-PET data with the structural MRI closest in time. In each subject’s native MRI space, tau-PET SUVR images were generated by normalizing mean tau-PET uptake to an inferior cerebellar gray matter defined by the spatially unbiased atlas template of the human cerebellum^88^. The tau data used in this study is corrected for partial volume effects using the geometric transfer matrix approach^89^.

From these imaging modalities, we extracted key imaging biomarkers, including hippocampal volume from MRI, Centiloid scale from Aβ-PET^90^ representing global cortical amyloid burden, and SUVR in the neocortex from tau-PET. Hippocampal volume was normalized to intracranial volume to account for individual differences in brain size. We also calculated the mean FDG-PET SUVR as an index of cortical metabolism by averaging regional SUVRs across the angular gyrus, temporal cortex, and posterior cingulate, regions known to exhibit early hypometabolism in AD. These imaging biomarkers were selected to provide a comprehensive representation of neurodegeneration, amyloid burden, and tau pathology, allowing us to investigate the associations between changes in these biomarkers and CSF proteomics profiles.

### Cognitive and functional (dementia severity) assessments

ADNI collects a broad set of instruments across cognition, function, global staging, and neuropsychiatric symptoms. Within this broader testing suite, we emphasize two widely used endpoints. ADAS-Cog11 serves as the cognitive measure: an 11-item scale covering memory, language, praxis, orientation, and comprehension/word-finding (e.g., word recall/recognition, naming, following commands, constructional and ideational praxis). Total scores range 0-70 (higher=worse). CDR-SB serves as the functional disease-severity/progression measure: a clinician-rated interview across six domains, three cognitive (memory, orientation, judgment/problem solving) and three functional (community affairs, home/hobbies, personal care), each scored 0-3 and summed to 0-18 (higher=worse). CDR-SB is particularly sensitive to early symptomatic change, while ADAS-Cog11 captures multi-domain cognitive decline. Both instruments are extensively validated and commonly used in trials and observational studies^56,57^.

### Statistical analysis and machine learning

#### Differential abundance analysis for key inflection points

Linear models were used to identify differentially abundant proteins (DAPs) associated with onset and progression. To account for multiple hypothesis testing, we applied the Benjamini-Hochberg false discovery rate (FDR) method to adjust p-values. The first set of comparisons focused on DAPs related to key inflection points, including conversion from Aβ-negative to Aβ-positive status, from asymptomatic to symptomatic stage (MCI [due-to-AD] or AD Dementia), and finally conversion from MCI [due-to-AD] to AD Dementia in already symptomatic individuals. Converted and non-converted participants were modelled as binary variables. For pathological and symptomatic conversion, all participants were included regardless of time to conversion or the maximum time of follow-up, whereas, for conversion to AD Dementia, given the number of sufficient samples, we restricted the analysis to 3-years cutoff and the participants who converted within 3-years or did not convert and had at least one follow-up timepoint after 3-years to confirm persistent MCI were included.

### Differential abundance analysis for longitudinal disease trajectories

For defining longitudinal disease trajectories across the clinical measures (CDR-SB, ADAS-Cog11) and neuroimaging biomarkers (tau-PET SUVR, FDG-PET SUVR, and hippocampal volume), participants having at-least 3 timepoints and data available for at-least 24 months were leveraged, except tau-PET and FDG-PET SUVR where 12-months cutoff were used and only two timepoints were required for calculation of longitudinal disease trajectories. All cognitive trajectories 4SD above or below the mean were removed given the inherent variability associated with longitudinal trajectories calculated from small number of visits. Rate of change of each outcome measure was calculated for each participant in R using the lme4 package^91^. Time was indexed as months from baseline and observations were grouped by participant ID. For each continuous outcome, we fit linear mixed-effects models with participant-specific intercepts and slopes and extracted individual rates of change for downstream analyses; participants were included per outcome when they had sufficient repeated measures for that particular outcome. Next, linear models were used to identify DAPs associated with the rate of change in each outcome. To account for multiple hypothesis testing, we applied the Benjamini-Hochberg method to adjust p-values using the FDR.

### Brain network module association to onset and progression in AD

In our prior work, we mapped 44 protein co-expression modules through deep, multilayer proteomics across more than 1,000 dorsolateral prefrontal cortex samples. Nearly half of these modules, including several strongly linked to AD traits, were absent from RNA networks, highlighting the distinct proteopathic architecture of AD. To connect these brain-derived network modules (M1-M44)^17^ with disease onset and progression, we evaluated CSF proteins associated with transitions as well as progression on clinical measures and neuroimaging biomarkers. Gene products along with corresponding p-values were tested for enrichment within brain proteome modules using a permutation-based framework (10,000 permutations) implemented in R. Exact p values were calculated using the ‘permp’ function from the statmod package. To quantify module-specific enrichment, we calculated Z scores by comparing the average p value of genes within each module to the distribution of mean p-values from 10,000 random permutations. The Z score was computed as the difference between the observed and permuted mean p-values, normalized by the standard deviation of the permuted distribution. This approach mirrors the enrichment analysis applied to MAGMA-derived gene-level statistics for genome-wide association study, as described in previous work^17^ (available at https://www.github.com/edammer/MAGMA.SPA).

### Subtype and Stage Inference (SuStaIn) in asymptomatic AD participants

To infer the temporal sequence of biological alterations accompanying progression from asymptomatic to symptomatic stage, we applied Subtype and Stage Inference (SuStaIn) clustering^45^ to cross-sectional CSF protein abundances, restricted to top 10 proteins showing significant differences between converters and non-converters. SuStaIn models disease progression within cross-sectional data as a probabilistic cascade of biomarker transitions, assigning individuals to subtypes and disease stages based on their molecular profiles. Because this analysis focused exclusively on converters, a single subtype was modeled while staging was performed across these participants to capture within-group progression. Model fitting was implemented using *pySuStaIn*. The inferred sequence of protein alterations was compared against a prior trajectory derived from targeted MS in the DIAN-TU cohort^42^ to assess reproducibility across cohorts and platforms. This analysis enabled evaluation of whether the DIAN-TU–based SuStaIn framework, originally established in autosomal-dominant AD, captures analogous protein cascades within the sporadic converter group.

### Modeling protein associations with time to event (pathologic or symptom onset)

We assessed protein-time associations by aligning CSF protein abundances to each participant’s time-to-event, defined as years between baseline CSF collection and either symptom onset or pathological onset (year 0 corresponding to the first symptomatic visit or first pathological conversion). For each outcome, linear regression models were fit at the group level with time-to-event as the sole predictor, and proteins were ranked by the strength of association. The top 10 proteins with the smallest p-values were selected for visualization. Because this analysis was cross-sectional rather than within-subject, fitted slopes represent population-level associations between protein abundance and proximity to onset. For display, protein abundances were plotted against time-to-event with corresponding regression lines for both symptom-onset and pathological-onset analyses.

### Defining participant groups for predictive modelling

We assessed the predictive capability of CSF proteins for both disease onset and longitudinal trajectories. Machine learning models were developed for each outcome, ensuring robust feature selection and classification. To capture inflection points, we analyzed three longitudinal transition groups within ADNI, each defined by baseline status and subsequent clinical or biological change. Amyloid onset was evaluated in 309 participants who were Aβ-negative by both Aβ42/pTau181 ratio^28^ and Aβ-PET and had longitudinal Aβ-PET scans to track conversion, among whom 271 remained Aβ-negative (Aβ-) over an average of 4.85 years, while 48 converted to Aβ-positivity (cAβ+) within 4.34 years. Symptomatic onset was examined in 100 participants who were asymptomatic at baseline (Aβ-positive by Aβ42/pTau181 ratio^28^ but cognitively unimpaired), of whom 64 remained stable (sAsym) and 34 progressed to MCI [due-to-AD] or AD dementia (cAsym). Clinical progression was assessed in 186 individuals diagnosed with MCI [due-to-AD] at baseline, including 61 who remained stable for at least 3 years (sMCI) and 125 who converted to AD Dementia within a 3-year timeframe (cMCI). For longitudinal disease trajectories, participants were stratified into slow and fast progressor groups based on the median slope of each outcome, yielding balanced subsets for classification: ADAS-Cog11 (n=406, slow=203, fast=203), CDR-SB (n=414, slow=207, fast=207), hippocampal volume (n=290, slow=145, fast=145), FDG-PET SUVR (n=241, slow=121, fast=120), and tau-PET SUVR (n=83, slow=42, fast=41). This categorization enabled us to model distinct phenotypic differences between slow and fast progressors.

### Predictive modelling using machine learning

To identify the most informative proteins distinguishing different groups of participants, we employed a two-step approach. First, feature selection was performed using the least absolute shrinkage and selection operator (LASSO)^37^ across 100 bootstrap iterations. Proteins selected in ≥90% of bootstraps were retained as robust stage-specific discriminative panels. Next, classification models were developed using random forest^36^ classification model, which are ensemble methods that combine many decision trees, providing strong predictive accuracy and robustness against overfitting^36^. They are especially effective for biomedical datasets because they naturally capture complex, non-linear relationships, handle high-dimensional features, and yield interpretable feature importance measures useful for biomarker discovery. Separate feature selection and random forest classification models were trained for each clinical outcome and neuroimaging measures. The dataset in each experiment was split into 80% training and 20% test sets, except the case of tau-PET SUVR where training/test data was split into 70-30% as data was limited and 80-20% split led to very small number of test samples. All feature selection and hyperparameter tuning (e.g., number of trees, depth of trees) were conducted exclusively within the training data using 5-fold cross-validation and grid search. The independent test set was held out entirely during model development and used solely to evaluate final performance. Machine learning analyses were conducted in Python (version 3.9), using libraries such as scikit-learn for model development and evaluation. Agreement between observed versus predicted group labels was assessed using area-under-the-receiver-operating-characteristic (ROC) curve. Optimal thresholds along the ROC were defined using the Youden index^92^. ROC curve analyses were performed in R using the pROC package to assess model performance and compute the area under the curve (AUC). Feature selection, model training, and validation on an independent test set were performed separately for each of the outcomes. Only proteins with data available for more than 95% of participants were included in machine learning. Remaining missing values were imputed using k-nearest neighbors (KNN) imputation, implemented via the VIM package in R. The reason behind selecting a panel before the classification was that we wanted to fix the panel and test it across 100 runs rather than selecting a new set of proteins on the training data in each iteration, however, similar results were obtained when the later approach was adopted.

We compared the performance of selected CSF protein panels with canonical CSF biomarkers, demographics, and clinical outcomes using a non-parametric permutation method. The null hypothesis assumed no difference in predictive performance between selected protein panels and conventional biomarkers. We first determined the actual performance difference between these models. Next, we randomly shuffled the assignment of CSF peptides and biomarkers for each participant and recalculated the performance difference. Statistical significance was determined using 100 permutations. The 100 permutations on protein panels and conventional biomarkers were performed in sync, making sure the same permuted training and test participants in each iteration.

In a smaller subset of participants with tau-PET and CSF proteomics both available at baseline, we compared the performance obtained using proteomics and tau-PET. For tau-PET, regional SUVR values from 68 FreeSurfer regions were used as input to train the model. Comparison with tau-PET was conducted only for ADAS-Cog11, CDR-SB, and hippocampal volume, where sufficient sample sizes were available.

## Data Availability

Raw mass spectrometry data and pre- and post-processed plasma protein abundance data and case traits related to this manuscript are available at https://www.synapse.org/Synapse:syn59804727 on the AMP-AD Knowledge Portal, which is a platform for accessing data, analyses and tools generated by the AMP-AD Target Discovery Program and other programs supported by the National Institute on Aging to enable open-science practices and accelerate translational learning. The data, analyses and tools are shared early in the research cycle without a publication embargo on secondary use. ADNI data are available on https://adni.loni.usc.edu/.

## Data Availability

Raw mass spectrometry data and pre- and post-processed plasma protein abundance data and case traits related to this manuscript are available at
https://www.synapse.org/Synapse:syn69962262. The AMP-AD Knowledge Portal is a platform for accessing data, analyses and tools generated by the AMP-AD Target Discovery Program and other programs supported by the National Institute on Aging to enable open-science practices and accelerate translational learning. The data, analyses and tools are shared early in the research cycle without a publication embargo on secondary use. Data are available for general research use according to the following requirements for data access and data attribution in the ADNI Biofluid Biomarker Core.

## Acknowledgements Section

This study was supported by the following National Institutes of Health (NIH) funding mechanisms: U01AG061357 (A.I.L. and N.T.S.), RF1AG062181 (N.T.S.), P30AG066511 (A.I.L.), RF1NS139948 (N.T.S.), and R01AG075820 (N.T.S.); as well as the Foundation for the National Institutes of Health (FNIH) AMP-AD 2.0 grant and a grant from the Alzheimer’s Association (ABA-22-974673). Data collection and sharing for this project was funded by the Alzheimer’s Disease Neuroimaging Initiative (ADNI) (National Institutes of Health Grant U01 AG024904) and DOD ADNI (Department of Defense award number W81XWH-12-2-0012). ADNI is funded by the National Institute on Aging, the National Institute of Biomedical Imaging and Bioengineering, and through generous contributions from the following: AbbVie, Alzheimer’s Association; Alzheimer’s Drug Discovery Foundation; Araclon Biotech; BioClinica, Inc.; Biogen; Bristol-Myers Squibb Company; CereSpir, Inc.; Cogstate; Eisai Inc.; Elan Pharmaceuticals, Inc.; Eli Lilly and Company; EuroImmun; F. Hoffmann-La Roche Ltd and its affiliated company Genentech, Inc.; Fujirebio; GE Healthcare; IXICO Ltd.; Janssen Alzheimer Immunotherapy Research & Development, LLC.; Johnson & Johnson Pharmaceutical Research & Development LLC.; Lumosity; Lundbeck; Merck & Co., Inc.; Meso Scale Diagnostics, LLC.; NeuroRx Research; Neurotrack Technologies; Novartis Pharmaceuticals Corporation; Pfizer Inc.; Piramal Imaging; Servier; Takeda Pharmaceutical Company; and Transition Therapeutics. The Canadian Institutes of Health Research is providing funds to support ADNI clinical sites in Canada. Private sector contributions are facilitated by the Foundation for the National Institutes of Health (www.fnih.org). The grantee organization is the Northern California Institute for Research and Education, and the study is coordinated by the Alzheimer’s Therapeutic Research Institute at the University of Southern California. ADNI data are disseminated by the Laboratory for Neuro Imaging at the University of Southern California. We also thank Dr. Vince Calhoun, Professor and Director of the Tri-Institutional Center for Translational Research in Neuroimaging and Data Science (TReNDS) at Georgia State University, Georgia Tech, and Emory University, for his insightful feedback.

## Disclosures

A.I.L. serves as a consultant to Cognito, Asha Therapeutics, NextSense and Cognition Therapeutics. N.T.S has consulted for AbbVie, Eisai, Trace Neuroscience and Arrowhead Pharmaceuticals. D.M.D., A.I.L., and N.T.S. are co-founders, employees, consultants, and/or shareholders of EmTheraPro. D.M.D. and N.T.S. are co-founders of Arc proteomics. N.T.S. is a co-founder of Stitch-Rx.

## Author Contributions

Conceptualization: S.R. and A.I.L; Formal analysis: S.R., E.B.D., A.S., N.T.S., and A.I.L.; Funding Acquisition: A.I.L., and N.T.S.; Investigation: S.R., E.B.D., A.S., N.T.S., and A.I.L.; Methodology: S.R., E.B.D. N.T.S., and A.I.L.; Project administration: S.R., A.I.L; Resources: S.R., E.B.D., J.J.L., E.C.B.J., N.T.S., and A.I.L.; Software: S.R., E.B.D., N.T.S., and A.I.L.; Supervision: A.I.L., N.T.S; Visualization: S.R., N.T.S. and A.I.L.; Writing-original draft: S.R., N.T.S., A.I.L.; Writing-review & editing: S.R., E.B.D., N.T.S, A.I.L; All authors read and approved the final manuscript.

## Supplementary Figures

**Fig. S1.**
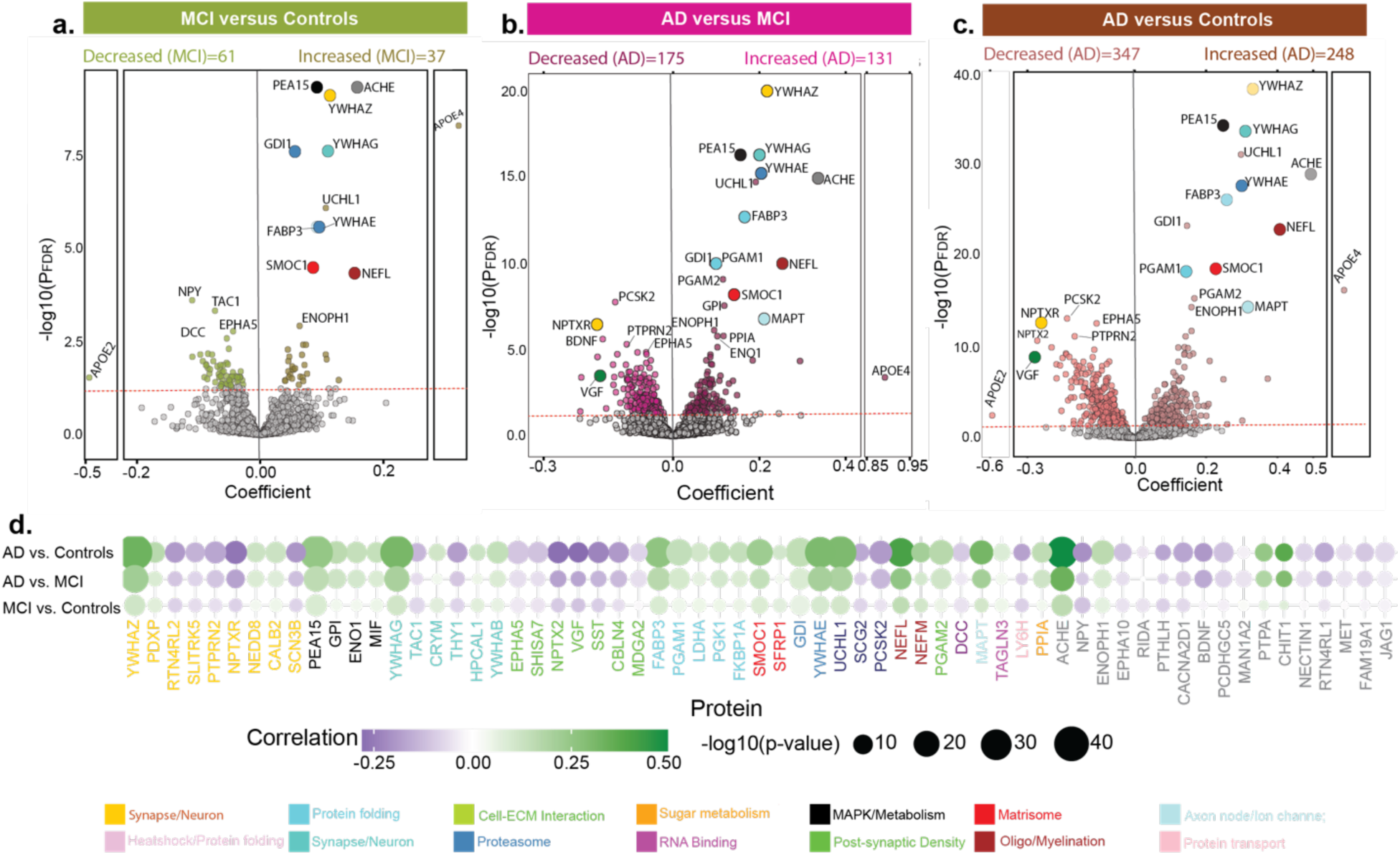
Differential abundance of CSF proteins across clinically diagnosed groups in ADNI. ***a-c,*** Volcano plots showing differential proteins abundance between ***a,*** MCI versus Controls (Increased: 37, Decreased: 61), ***b,*** AD versus MCI (Increased: 131, Decreased: 175), and ***c,*** AD versus Controls (Increased: 248, Decreased: 347) (Controls: n=377, MCI: n=563, AD: n=164). The dashed red line indicates the significance threshold at α = 0.05 after FDR correction. Top proteins are labeled for clarity, and selected proteins of interest are zoomed in. The x axis represents the regression coefficient while the y axis shows the -log10 of the FDR-corrected p-value calculated for each protein. ***d,*** Heatmap of regression coefficients showing associations between CSF proteomic profiles and clinical diagnosis. CSF proteins are labeled by their respective gene symbols, and the strength and direction of correlation are indicated by the purple-to-green color scale. The top 30 proteins were selected (based on P value) from each category, and their union is displayed in the heatmaps. Colors are assigned to individual protein names based on their membership in the brain-derived modules^17^, consistent across both the volcano plots and heatmaps. In the heatmap, proteins shown in gray were either not present in the prior network study or were not assigned to any module^17^.

**Fig. S2.**
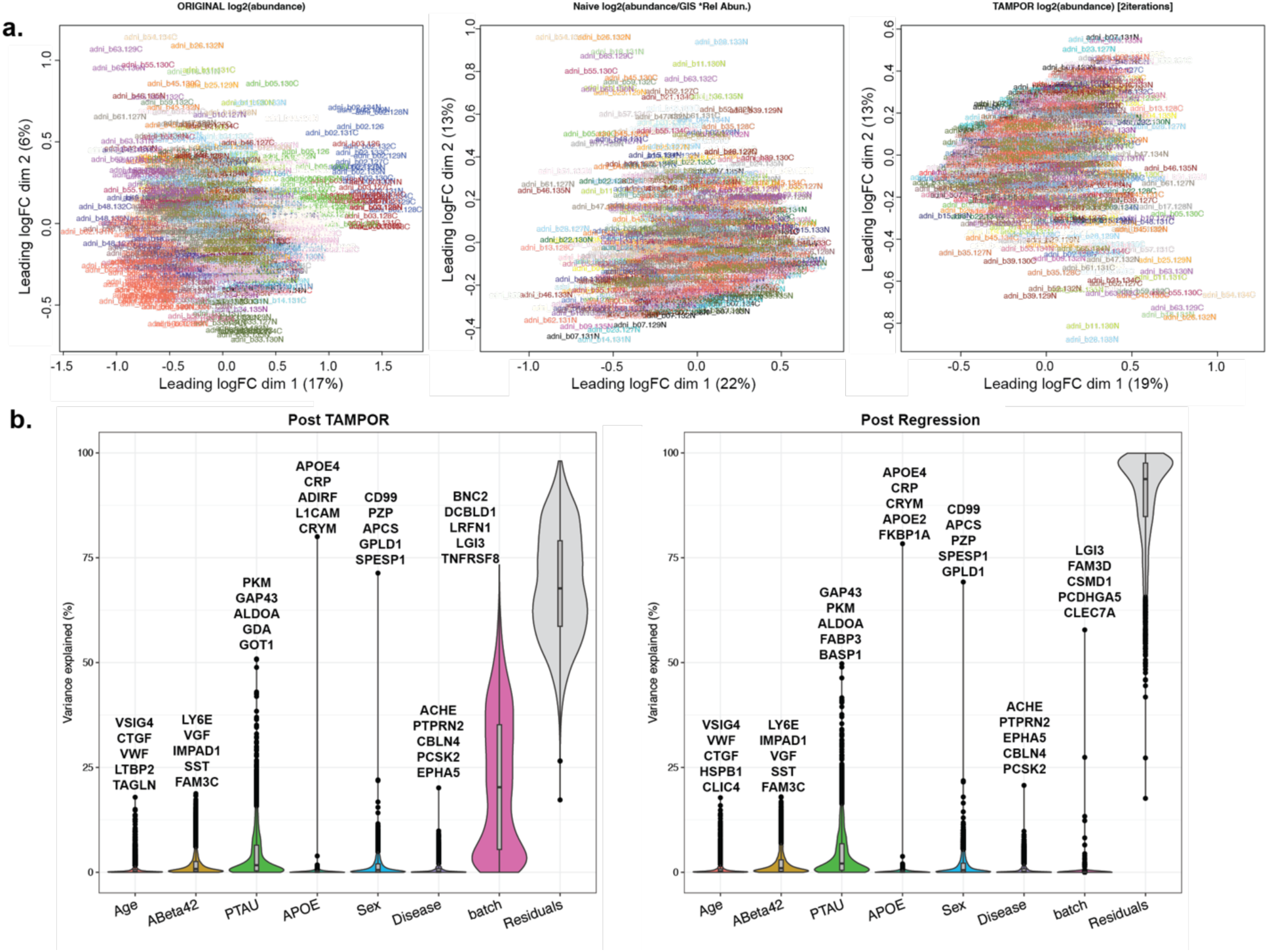
Quality control of the ADNI CSF proteome. **a,** Multidimensional scaling (MDS) illustrating TMT-MS batch correction. Log_2_ abundance, log_2_ abundance divided by the global internal standard (GIS), and TAMPOR are shown. **b,** Variance partition plots were used to visualize the percent variance of each protein in the dataset co-varying with batch, age and sex. The matrix was subjected to bootstrap regression (right) to remove variance due to batch.

**Fig. S3.**
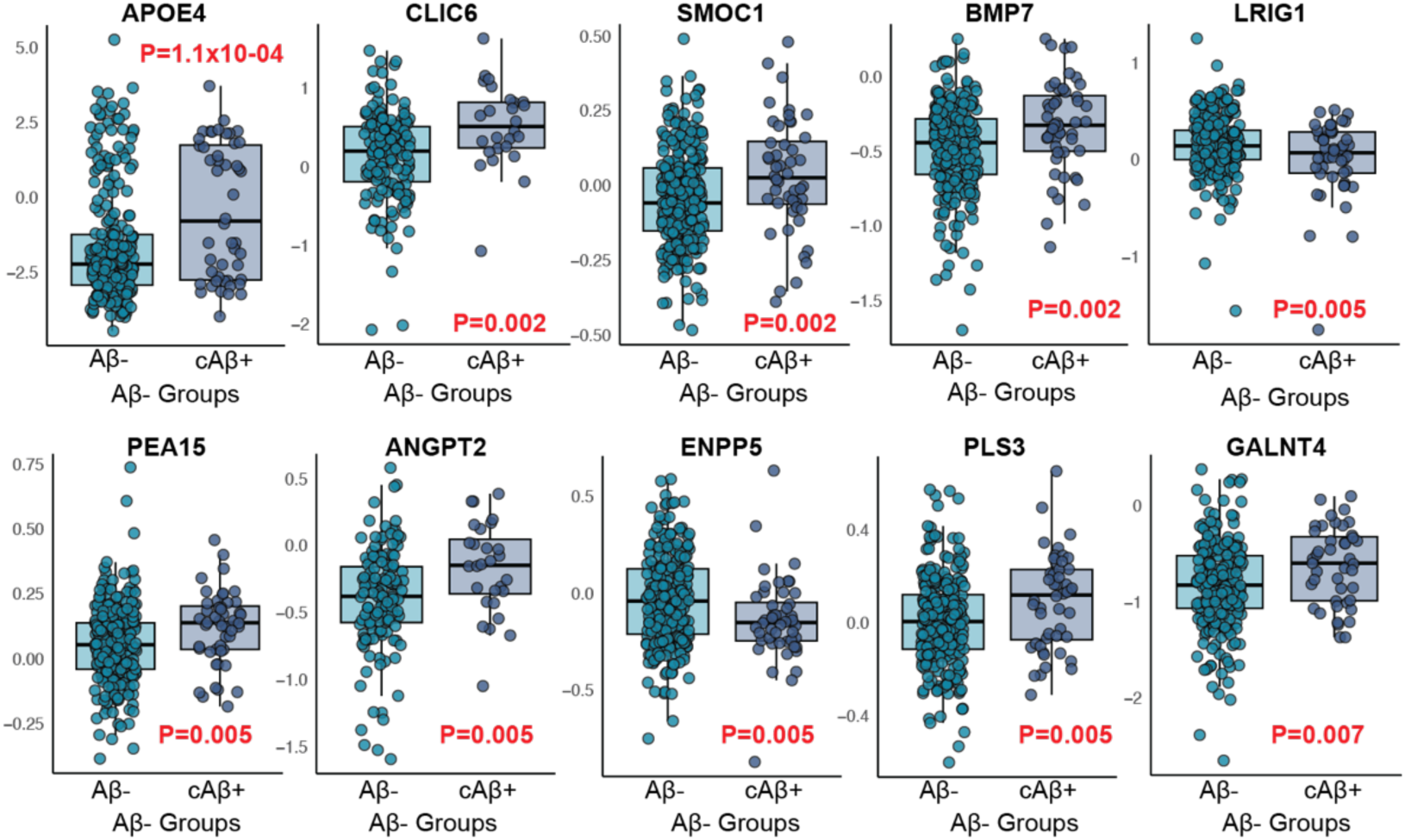
Proteins significantly associated with pathological onset. Box plots show abundance of top 10 proteins identified by differential abundance analysis between Aβ-participants who remained stable (Aβ-) and those who converted to Aβ+ (cAβ+). P values for each protein were obtained using a linear regression model. Box plots display the median and interquartile range (25th-75th percentile), with whiskers extending up to 1.5 times the interquartile range. Data points represent individual participants.

**Fig. S4.**
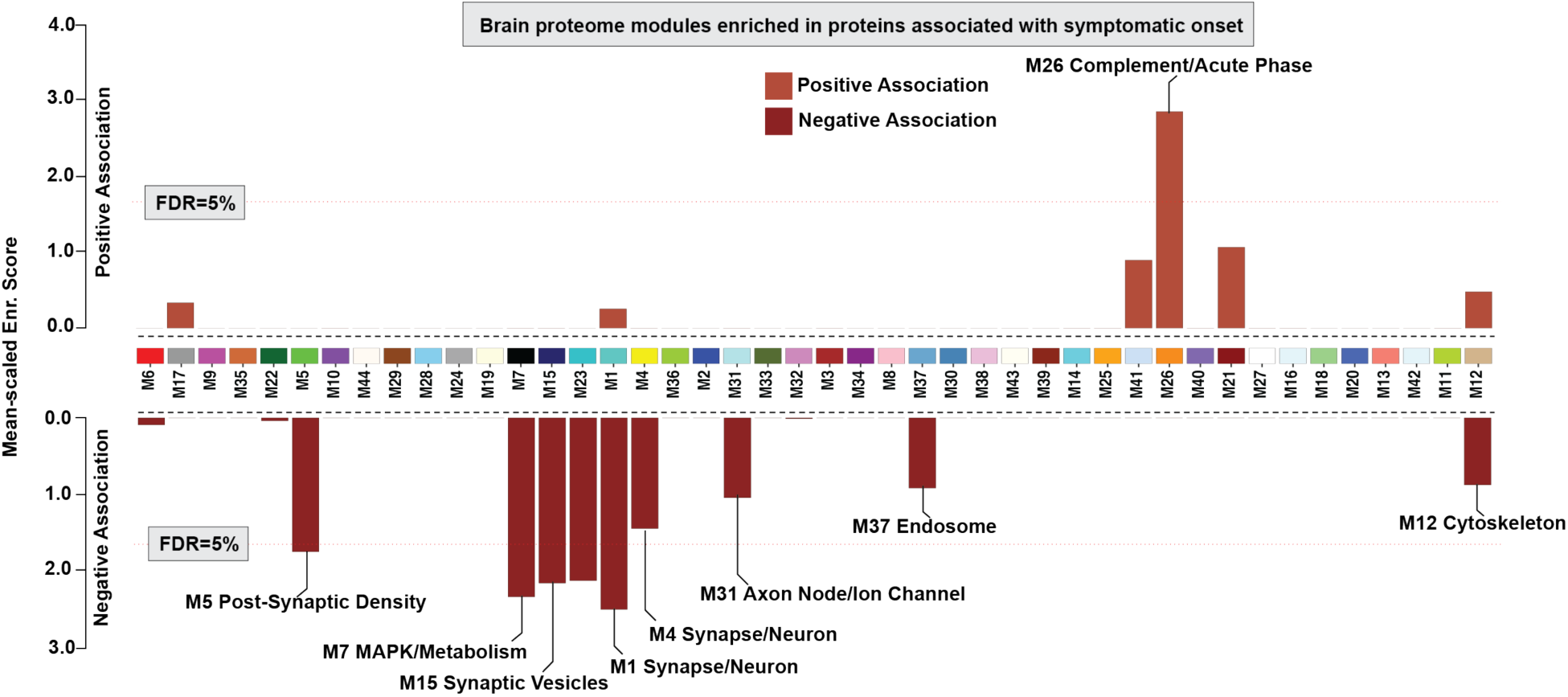
Brain network modules (M5, M7, M15, M1, and M26) enriched for CSF biomarkers of symptomatic onset. CSF proteins associated with symptomatic onset (P<0.05) were mapped onto brain proteome network modules ^17^. The horizontal red dotted line marks the 5% false discovery rate (FDR) threshold from permutation testing, above which the enrichment was considered significant. Modules with significant enrichment or biologically relevant are labeled, with proteins positively and negatively associated shown separately to highlight directionality. Unlabeled but significant modules lacked defined annotations in the original network study.

**Fig. S5.**
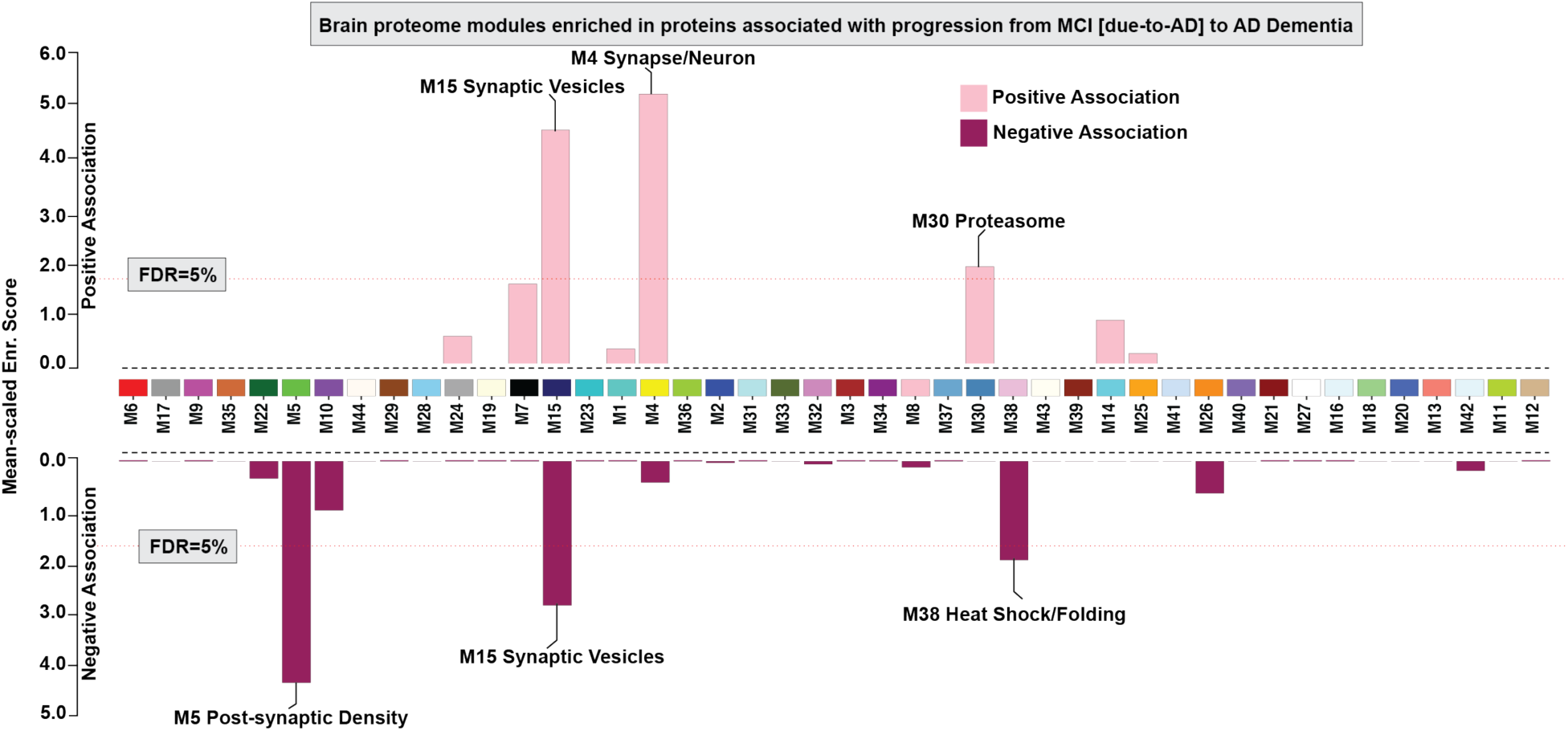
Brain network modules (M5, M15, M4, M30, and M38) enriched for CSF biomarkers of disease progression from MCI [due-to-AD] to AD Dementia. CSF proteins associated with disease progression (P<0.05) were mapped onto brain proteome network modules^17^. The horizontal red dotted line marks the 5% false discovery rate (FDR) threshold from permutation testing, above which the enrichment was considered significant. Modules with significant enrichment are labeled, with proteins positively and negatively associated shown separately to highlight directionality. Some proteins negatively and others positively associated with progression were enriched in module M15.

**Fig. S6.**
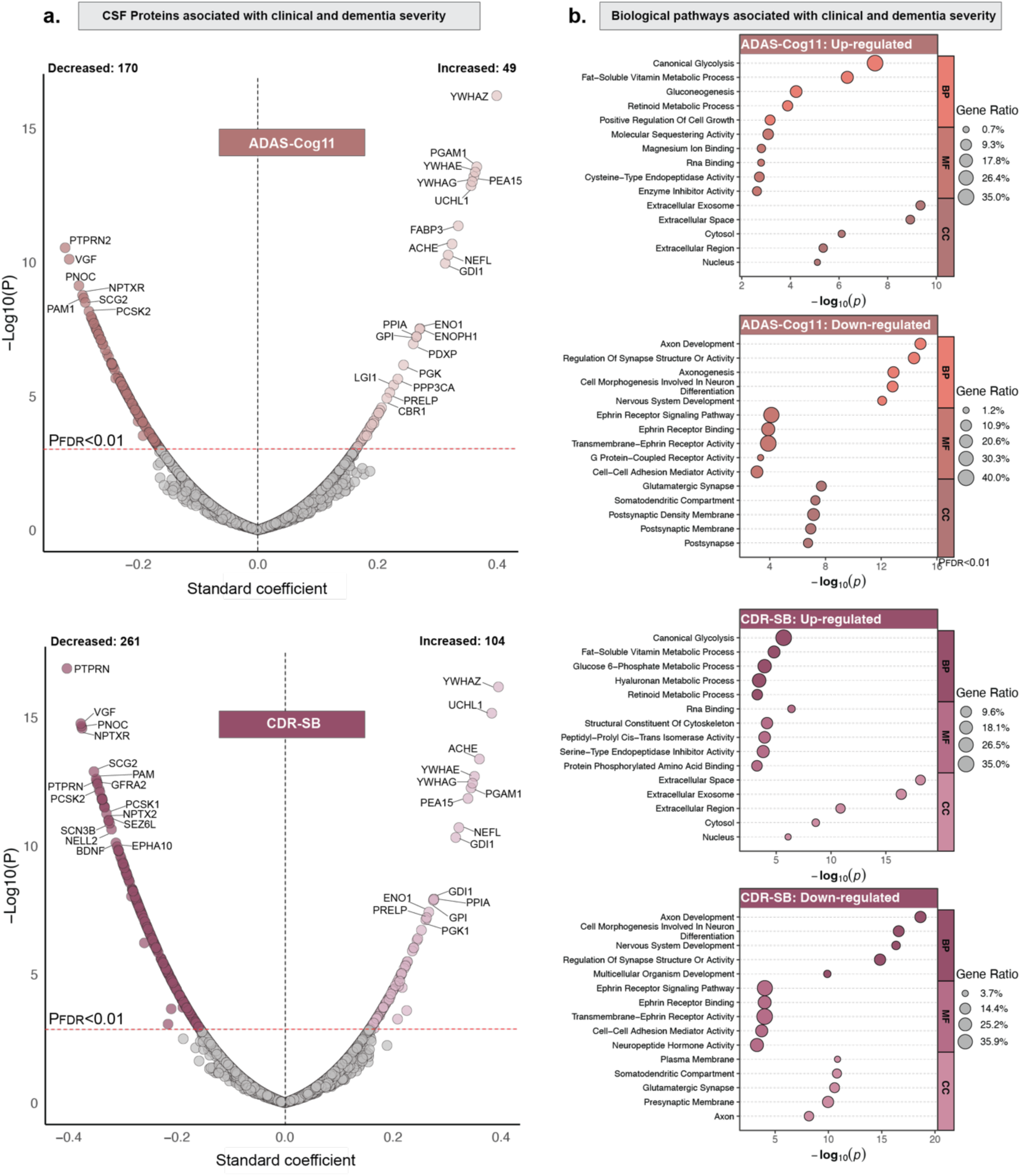
Biological processes underlying the rate of change in clinical and dementia severity measures. ***a,*** Volcano plots depict differentially expressed proteins associated with the rate of change in cognitive (ADAS-Cog11; Increased: 49; Decreased: 170) and dementia severity measures (CDR-SB; Increased: 104; Decreased: 261). Linear mixed-effects models were used to estimate subject-specific rates of change separately for all outcomes. P values were adjusted for FDR, and the red line represents the threshold of P_FDR_<0.01, above which proteins were considered significant. The x-axis shows the regression coefficient, and the y-axis indicates the -log10(P) for all proteins. Only the top proteins are labeled for legibility on the volcano plots. ***b,*** Gene ontology analysis was performed to identify biological processes associated with the rate of change in cognitive (ADAS-Cog11) and dementia severity measures (CDR-SB) using proteins significantly associated with each clinical measure at P<0.05, as shown in Panels **(a)** and Fig. 5. Upregulated and downregulated pathways are separately shown to highlight the directionality of the associations. Sample sizes were: ADAS-Cog11 (N = 406), and CDR-SB (N = 414).

**Fig. S7.**
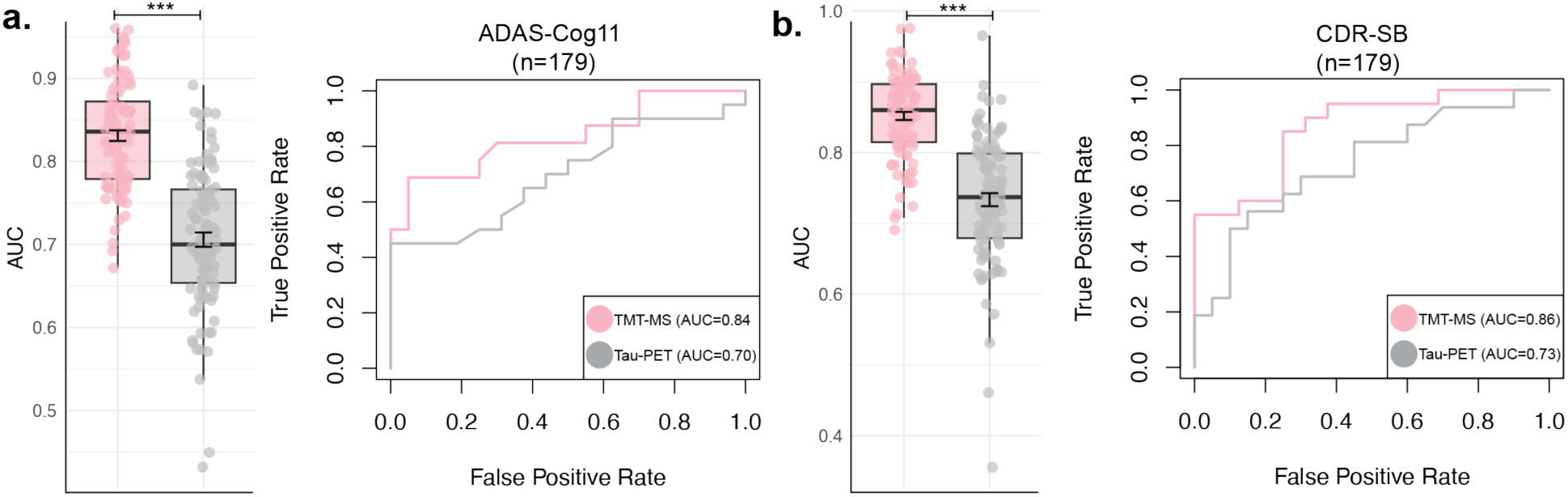
CSF proteomics predicts longitudinal trajectories of clinical and dementia severity measures, beyond tau-PET imaging biomarker. Longitudinal trajectories of participants in terms of cognitive (ADAS-Cog11) and dementia severity (CDR-SB) measures were calculated leveraging participants having at-least 3 timepoints and data available for at least 24 months. Participants were divided into fast-versus stable/slow progressors based on median cutoff slope. Bar plot show the AUC of the models across 100 permuted runs in predicting stable/slow- and fast-progressors in longitudinal disease trajectories and the ROC curve showing the classification performance pertaining to median AUC is shown for **a.** ADAS-Cog11 and **b.** CDR-SB, using the following predictors: 1) the CSF proteins (“TMT-MS”), and 2) regional free-surfer based tau-PET SUVR. (*p < 0.05, **p < 0.01, ***p < 0.001). Sample sizes are: ADAS-Cog11: n=179 and CDR-SB: n = 179.

**Fig. S8.**
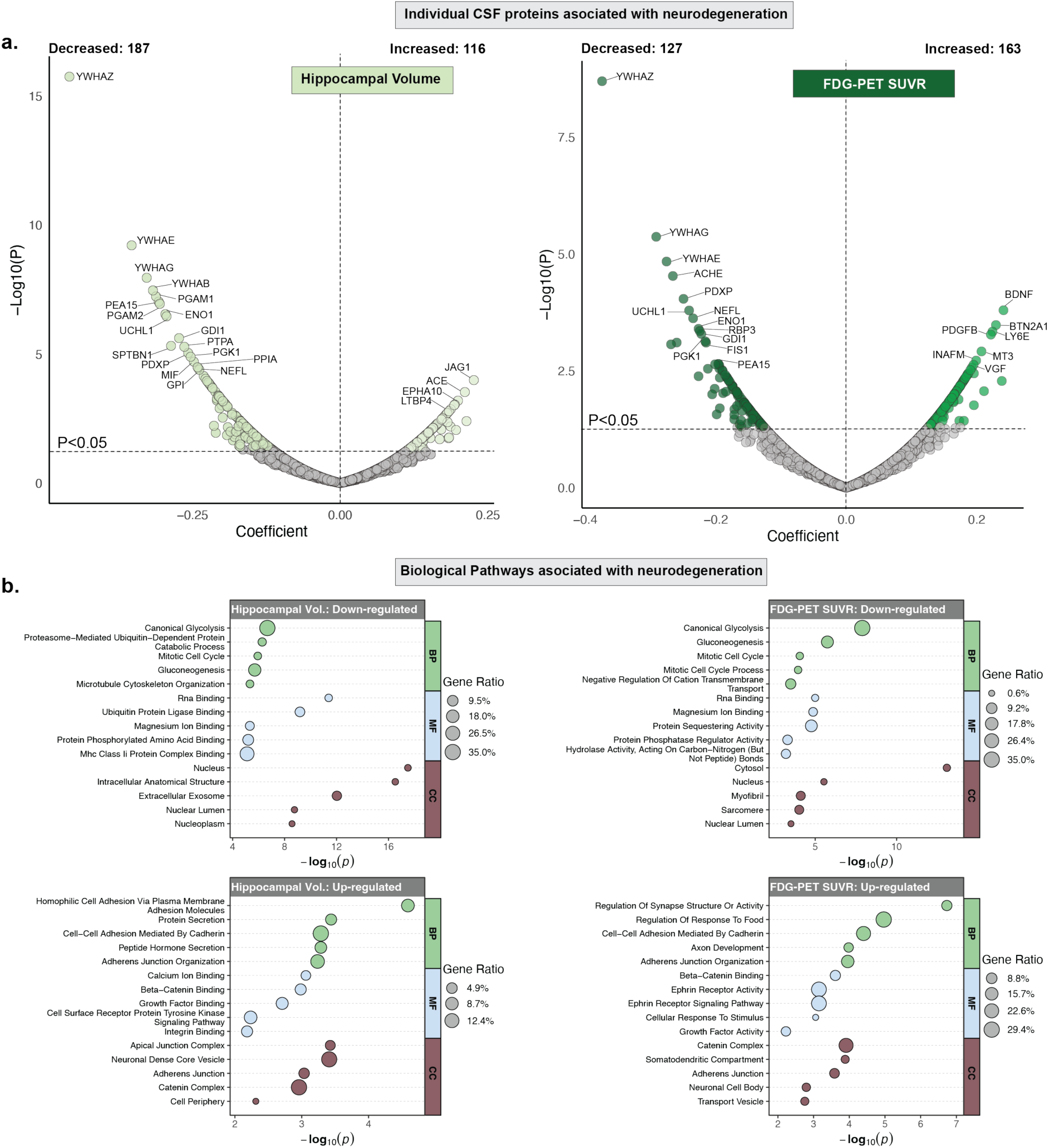
Biological processes underlying the rate of change in neuroimaging-based neurodegeneration. ***a,*** Volcano plots depict differentially abundant proteins associated with the rate of change in neurodegeneration-related measures, including hippocampal volume, normalized by intracranial volume (Increased: 116; Decreased: 187) and FDG-PET SUVR, defined as the mean of angular, temporal, and posterior cingulate regions (Increased: 163; Decreased: 127). Linear mixed-effects models were used to estimate subject-specific rates of change separately for all outcomes. The black line indicates the threshold of P<0.05, above which proteins were considered significant. The x-axis shows the regression coefficient, and the y-axis indicates the -log10(P) for all proteins. Only the top proteins are labeled for legibility on the volcano plots. ***b,*** Gene ontology analysis was performed to identify biological processes associated with the rate of change in hippocampal volume and FDG-PET SUVR using proteins significantly associated with each imaging biomarker, as shown in Panels ***(a)*** and Fig. 6. Upregulated and downregulated pathways are shown separately to highlight the directionality of associations. Sample sizes were: Hippocampal volume (N = 290) and FDG-PET SUVR (N = 241).

**Fig. S9.**
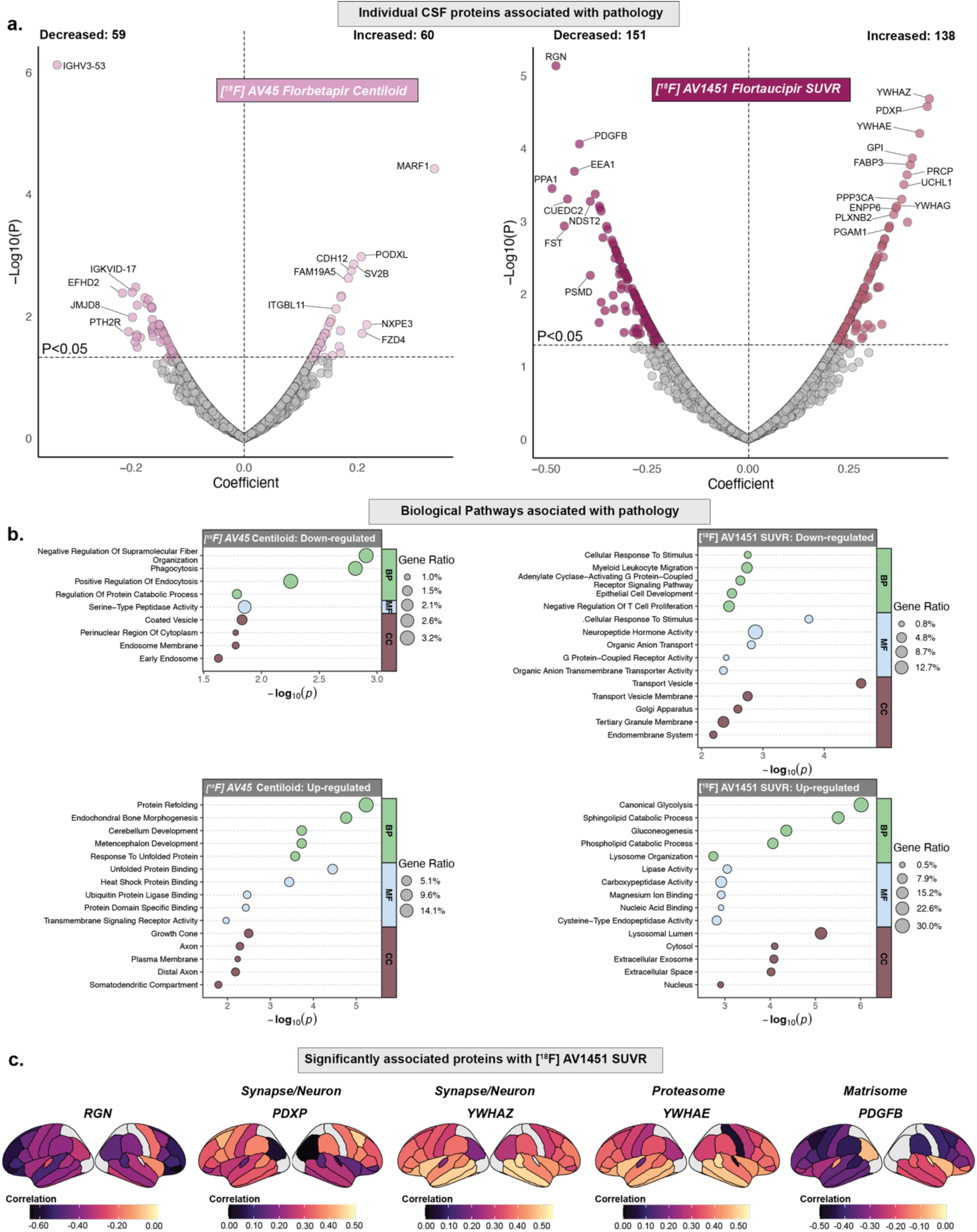
Biological processes underlying the rate of change in neuroimaging-based pathological accumulation. ***a,*** Volcano plots depict differentially expressed proteins associated with the rate of change in amyloid accumulation ([^18^F] AV45 PET Centiloid scale; Increased: 60; Decreased: 59) and neocortical tau accumulation ([^18^F] AV1451 PET SUVR; Increased: 138; Decreased: 151). Linear mixed-effects models were used to estimate subject-specific rates of change separately for all outcomes. The black line indicates the threshold of P<0.05, above which proteins were considered significant. The x-axis shows the regression coefficient, and the y-axis indicates the -log10(P) for all proteins. Only the top proteins are labeled for legibility on the volcano plots. ***b,*** Gene ontology analysis was performed to identify biological processes associated with the rate of change in amyloid (([^18^F] AV45 PET Centiloids scale) and tau accumulation ([^18^F] AV1451 PET SUVR) using the proteins significantly associated with each imaging phenotype, as shown in panel **a** and Fig. 6. Upregulated and downregulated pathways are separately shown to highlight the directionality of the associations. ***c,*** Surface maps show correlations between regional tau-PET SUVR accumulation and top-most selected five proteins from panel **a**. Sample sizes were: Aβ-PET Centiloids (N = 270), and [^18^F] AV1451 PET SUVR (N = 83).

**Fig. S10.**
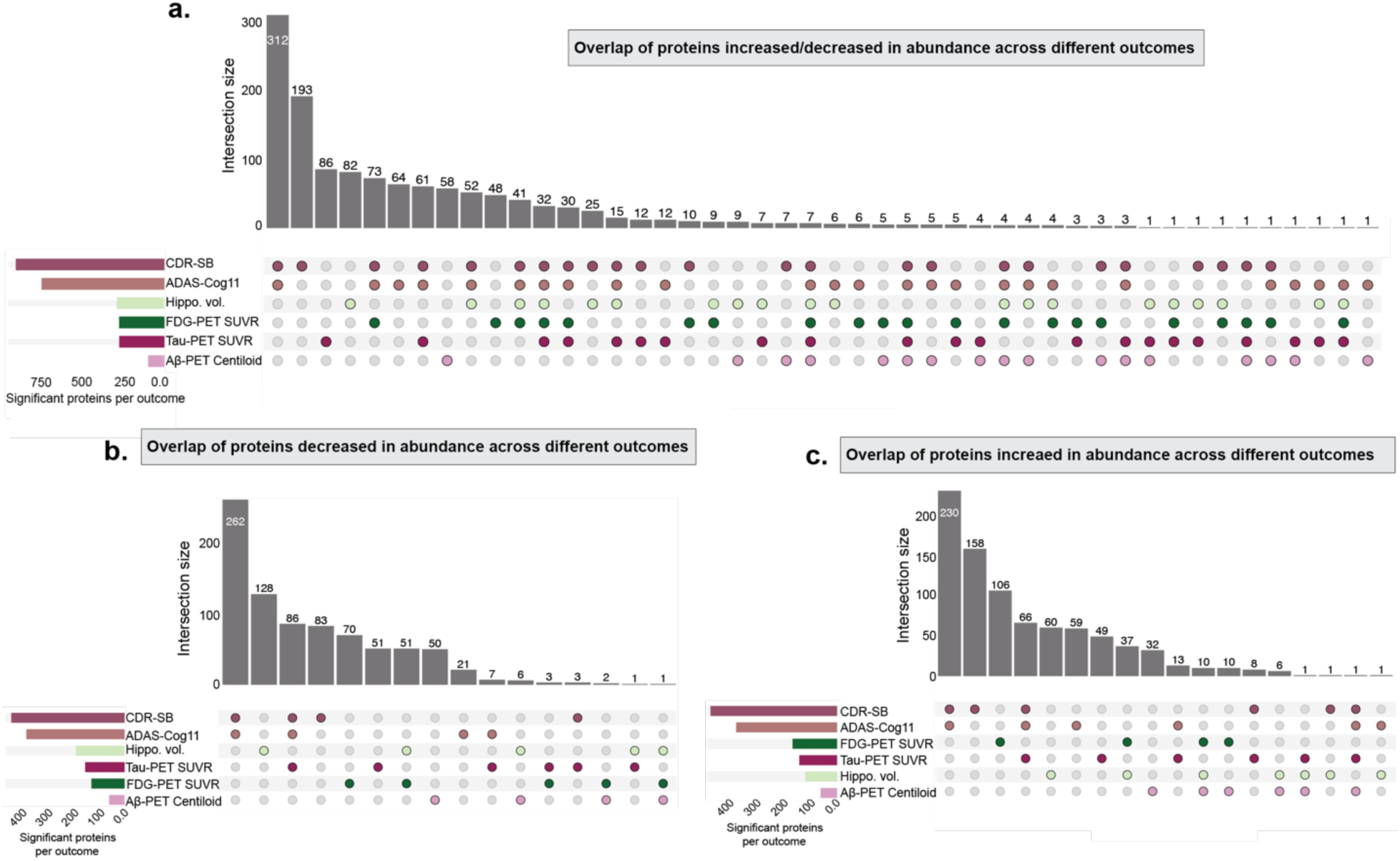
UpSet plots showing the overlap of CSF proteins significantly associated with cognitive and dementia severity outcomes, as well as with neurodegeneration- and pathology-related measures. ***a,*** All significant proteins (increased or decreased in abundance); ***b,*** Proteins showing decreased abundance. ***c,*** Proteins showing increased abundance. Only FDR-significant proteins (P < 0.05) were included in the overlap analysis.

